# Histology-guided MRI segmentation of brainstem nuclei critical to consciousness

**DOI:** 10.1101/2024.09.26.24314117

**Authors:** Mark Olchanyi, Jean Augustinack, Robin L. Haynes, Laura D. Lewis, Nicholas Cicero, Jian Li, Christophe Destrieux, Rebecca D. Folkerth, Hannah C. Kinney, Bruce Fischl, Emery N. Brown, Juan Eugenio Iglesias, Brian L. Edlow

**Author notes:** co-senior authors (Juan Eugenio Iglesias, Brian L. Edlow).

## Abstract

While substantial progress has been made in mapping the connectivity of cortical networks responsible for conscious awareness, neuroimaging analysis of subcortical arousal networks that modulate arousal (i.e., wakefulness) has been limited by a lack of a robust segmentation procedures for brainstem arousal nuclei. Automated segmentation of brainstem arousal nuclei is an essential step toward elucidating the physiology of arousal in human consciousness and the pathophysiology of disorders of consciousness. We created a probabilistic atlas of brainstem arousal nuclei built on diffusion MRI scans of five *ex vivo* human brain specimens scanned at 750 µm isotropic resolution. Labels of arousal nuclei used to generate the probabilistic atlas were manually annotated with reference to nucleus-specific immunostaining in two of the five brain specimens. We then developed a Bayesian segmentation algorithm that utilizes the probabilistic atlas as a generative model and automatically identifies brainstem arousal nuclei in a resolution- and contrast-agnostic manner. The segmentation method displayed high accuracy in both healthy and lesioned *in vivo* T1 MRI scans and high test-retest reliability across both T1 and T2 MRI contrasts. Finally, we show that the segmentation algorithm can detect volumetric changes and differences in magnetic susceptibility within brainstem arousal nuclei in Alzheimer’s disease and traumatic coma, respectively. We release the probabilistic atlas and Bayesian segmentation tool in FreeSurfer to advance the study of human consciousness and its disorders.

## 1 INTRODUCTION

Over the past two decades, connectivity studies of cortical networks have begun to reveal the structural and functional correlates of human cognition (Buckner & DiNicola, 2019; Glasser et al., 2016; Yeo et al., 2011). Observations about the spatial and temporal dynamics of cortical network connectivity (Fox et al., 2005; Horn et al., 2014; Sporns et al., 2005) have yielded insights into the neuroanatomic basis of language, memory, attention, emotion, and conscious awareness (Cole et al., 2014; Demertzi et al., 2019; Glasser et al., 2016; Medaglia et al., 2015). In parallel, network-based models of neuropsychiatric diseases have emerged (Fox, 2018), based on the observation that spatially disparate lesions can cause cognitive deficits and behavioral dysfunction by disrupting a shared network architecture (Bodien et al., 2017; Boes et al., 2015; Fischer et al., 2016; Snider et al., 2020).

Yet as cortical connectivity mapping has accelerated, progress in mapping the subcortical networks that modulate arousal (i.e., wakefulness) has lagged behind. Few studies have attempted to map the complex connectivity of brainstem networks (Beissner et al., 2014; Bianciardi et al., 2016; Edlow et al., 2012, 2016, 2024; Li et al., 2021; Sclocco et al., 2018). As a result, fundamental questions about the pathogenesis of a broad range of disorders of arousal, including coma (Edlow et al., 2021), sudden infant death syndrome (Kinney & Haynes, 2019), and post-COVID-19 fatigue (Huang et al., 2021) remain unanswered. This gap in knowledge is partly attributable to the lack of robust and automated methods for segmenting brainstem nodes of the ascending arousal network (AAN), which modulates arousal (Edlow et al., 2012; Parvizi, 2001; Valenza et al., 2019).

To date, brainstem segmentation methods have mainly focused on the brainstem as a whole. The brainstem is extracted by segmentation modules of most neuroimaging packages, such as “aseg” (Fischl et al., 2002) in FreeSurfer (Fischl, 2012) or FIRST (Patenaude et al., 2011) in FSL (Smith et al., 2004). The whole brainstem has also been targeted in multi-atlas segmentation methods (Heckemann et al., 2006), as well as specifically designed methods, such as (Bondiau et al., 2005) based on a single labeled template, or (Jiann-Der Lee et al., 2005; J. Lee et al., 2007) based on active contours.

Beyond whole-brainstem segmentation, several methods have segmented the brainstem into its three main neuroanatomic components – the medulla, pons and midbrain – based on manual procedures (Lechanoine et al., 2021), geometric rules (Nigro et al., 2014) or Bayesian methods (Ashburner & Friston, 2005; Iglesias, Van Leemput, et al., 2015; Lambert et al., 2013). However, automated segmentation techniques for individual brainstem nuclei beyond simple registration to a single labeled template (e.g., the Harvard AAN Atlas) (Edlow et al., 2012, 2024) have not yet been developed. Recently, several teams of investigators leveraged ultra-high resolution imaging datasets to provide anatomic atlases of brainstem nuclei (Adil et al., 2021; Bianciardi, 2021; Lechanoine et al., 2021), but these atlases do not segment all of the miniscule brainstem nuclei in the pontine and midbrain tegmentum that are critical to arousal and homeostasis.

Here, we develop a probabilistic brainstem AAN atlas from manual tracings made on *ex vivo* MRI data acquired at 750 µm resolution, guided by a supporting 200 µm MRI sequence and nucleus-specific immunostain data from two of the scanned human brain specimens. *Ex vivo* MRI provides substantial improvements in signal-to-noise ratio and spatial resolution over *in vivo* MRI by reducing motion and enabling long scanning times (Edlow et al., 2019; McNab et al., 2009; Yendiki et al., 2022). These high-resolution *ex vivo* images facilitate precise manual delineation of AAN structures, which in turn enables building an atlas with a superior level of detail. We then used the new probabilistic version of the Harvard AAN Atlas as the basis for creation of a companion automated algorithm that segments AAN brainstem arousal nuclei with *in vivo* MRI. The segmentation algorithm is based on Bayesian inference using generative models of brain MRI data (Ashburner & Friston, 2005; Pohl et al., 2006; Van Leemput et al., 1999; Wells et al., 1996). Because the modeling of intensities is unsupervised, this approach allows investigators to apply atlases built in *ex vivo* brain specimens to the segmentation of *in vivo* MRI scans (Iglesias, Augustinack, et al., 2015; Iglesias et al., 2018; Saygin et al., 2017). To demonstrate the translational potential of this automated segmentation tool, we apply it to a volumetric analysis of AAN brainstem nuclei in patients with Alzheimer’s disease (AD) and acute traumatic disorders of consciousness (DoC), as compared to healthy human subjects. We release the AAN automated segmentation tool as part of the FreeSurfer neuroimaging package to facilitate a broad range of potential applications in the study of human consciousness and its disorders. The tool can be found at surfer.nmr.mgh.harvard.edu/fswiki/AANSegment.

## 2 METHODS

### 2.1 Ex vivo brain specimen overview

We analyzed five human brain specimens: two using histological sectioning (S1, S2) and five using *ex vivo* MRI (S1, S2, S3, S4, S5). All brains were donated by individuals with no history of neurological or psychiatric disease and who died of non-neurological causes. Consent for brain donation and research was provided by surrogate decision-makers as part of Institutional Review Board protocols at Mass General Brigham (S1-S3) or at the Université de Tours (S4, S5). Postmortem examination of each brain specimen by a neuropathologist was grossly normal.

Brain specimens were extracted from the cranium and fixed in 10% formalin (S1-4) or 5% formalin (S5), for at least 23 months prior to *ex vivo* MRI. Immediately prior to scanning, all specimens were transferred to a fomblin solution (Solvay Specialty Polymers, Bollate, Italy), to reduce artifacts related to magnetic susceptibility, as previously described (Edlow et al., 2018). Specimen demographics, causes of death, and fixation parameters are summarized in Table 1.

**Table 1:** Demographics and details on post-mortem fixation for all *ex vivo* brain specimens. DVT: Deep Vein Thrombosis. HTN: Hypertension, DIC: Disseminated Intravascular Coagulation, PE: pulmonary embolism, DAD: Diffuse Alveolar Damage.

| Specimen number | Age | Sex | Medical history | Cause of death | Post-mortem fixation interval (h) | Fixative | Fixation-to-imaging duration (M) | Histology included |
| --- | --- | --- | --- | --- | --- | --- | --- | --- |
| 1 | 56-60 | F | Cecal adenocarcinoma (metastatic), DVT, depression | Septic shock | <24 | 10% formalin | 24 | yes |
| 2 | 61-65 | F | HTN, ovarian cancer (metastatic) | Septic shock | 72 | 10% formalin | 20 | yes |
| 3 | 46-50 | M | Depression, leukemia, Raynaud's phenomena, thromboembolism | DIC due to Hemophagocytic lymphohistiocytosis | <24 | 10% formalin | 93 | no |
| 4 | 56-60 | F | Breast cancer, ulcerative colitis | PE and/or DAD in setting of widely metastatic breast cancer | <24 | 10% formalin | 92 | no |
| 5 | 81-85 | M | Hyperlipidemia, chronic respiratory deficiency | Acute respiratory failure | 23 | 5% formalin | 50 | no |

### 2.2 Data acquisition

#### 2.2.1 Ex vivo MRI

Each *ex vivo* brain specimen (S1-S5) was scanned on a 7 Tesla (7T) Siemens Magnetom scanner (Siemens Healthineers, Erlangen, Germany) and a 3 Tesla (3T) Siemens Tim Trio scanner (Siemens Healthineers, Erlangen, Germany). The 7T Fast Low-Angle SHot (FLASH) sequence (Augustinack et al., 2005; Fischl et al., 2004) utilized the following parameters: TR = 40ms, TE = 14.2ms, flip angle = 20°, acquired at 200 µm isotropic spatial resolution. The 3T diffusion-weighted steady-state free procession (DWSSFP) sequence (McNab et al., 2009) utilized the following parameters: TR = 38ms, TE = 23ms, flip angle = 60°, with 90 diffusion-encoding directions and 12 low-b images acquired at 750 µm isotropic spatial resolution.

#### 2.2.2 Histology and immunostaining

The two specimens that underwent histological analysis (S1, S2) were sectioned and stained in the axial (i.e., transverse) plane using a standardized protocol, as previously described (Edlow et al., 2024). Briefly, the brainstem was dissected from the brain specimen via a transverse cut at the mesencephalic-diencephalic junction. Each brainstem was then separated into four blocks (medulla, caudal pons, rostral pons, and midbrain), which were embedded in paraffin. Serial sections were cut at 10 μm thickness from the paraffin-embedded blocks using a microtome (LEICA RM2255 microtome, Leica Microsystems, Buffalo Grove, IL, USA). Every 250 µm, a section was stained with hematoxylin and eosin, and counterstained with Luxol fast blue (H&E/LFB), for identification of cell bodies and myelin. Sections were then selected based on anatomic landmarks to identify brainstem arousal nuclei at the level of the rostral pons (pontine reticular formation, median raphe, locus coeruleus, parabrachial complex, laterodorsal tegmental nucleus), caudal midbrain (mesencephalic reticular formation, ventral tegmental area, pedunculotegmental nucleus, dorsal raphe, periaqueductal grey) and rostral midbrain (mesencephalic reticular formation, ventral tegmental area, and periaqueductal grey). Tyrosine hydroxylase immunostaining was used to identify the VTA and LC, tryptophan hydroxylase staining to identify the MnR and DR, and H&E/LFB to identify the PnO, LC, PBC, LDTg, PTg, and PAG. A summary of the brainstem arousal nuclei that were assessed, along with their immunostaining and histological characteristics, is provided in Table 2. All histological sections were previously digitized with the NanoZoomer S60 Digital Slide Scanner (Hamamatsu Photonics). Digitized slides underwent a custom whole-slide image processing pipeline for white balance correction and contrast enhancement. We converted all histological slides to JPEG2000 format and previously published them through the Biolucida platform for visualization.

**Table 2:** Description of AAN nuclei, including their primary neurotransmitter-specific cell bodies, histological staining, and corresponding locations for manual annotation.

| Brainstem arousal nucleus | Abbreviation | ROI color | Primary neurotransmitter | Histology and immunohistochemistry |
| --- | --- | --- | --- | --- |
| Dorsal Raphe | DR |  | Serotonin | Positive staining for anti-tryptophan hydroxylase |
| Median Raphe | MnR |  | Serotonin | Positive staining for anti-tryptophan hydroxylase |
| Locus Coeruleus | LC |  | Norepinephrine | Positive staining for tyrosine hydroxylase, and visible in H&E-LFB |
| Laterodorsal Tegmental Nucleus | LDTg |  | Acetylcholine | Annotated with H&E-LFB |
| Parabrachial Complex | PBC |  | Glutamate | Annotated with H&E-LFB |
| Pontis Oralis | PnO |  | Glutamate | Annotated with H&E-LFB |
| Midbrain Reticular Formation | mRt |  | Glutamate | Annotated with H&E-LFB |
| Pedunculotegmental nucleus | PTg |  | Acetylcholine | Annotated with H&E-LFB |
| Periaqueductal Grey | PAG |  | Multiple | Annotated with H&E-LFB |
| Ventral Tegmental Area | VTA |  | Dopamine | Positive staining tyrosine hydroxylase |

#### 2.2.3 In vivo MRI of traumatic brain injury patients

We acquired *in vivo* MRI data from 25 healthy control subjects and 18 patients with severe traumatic brain injury (TBI). The patients with severe TBI were scanned during the acute phase of injury in the intensive care unit as part of a previously published observational study (ClinicalTrials.gov: NCT03504709) at Massachusetts General Hospital (Edlow et al., 2017). Informed consent was obtained from the healthy control subjects and from surrogate decision makers for the patients with severe TBI, in accordance with a protocol approved by the Mass General Brigham Institutional Review Board. Healthy controls had no history of neurological, psychiatric, or medical diseases. Pertinent inclusion criteria for patients with acute severe TBI were: a Glasgow Coma Scale (GCS) (Teasdale & Jennett, 1974) score of less than or equal to 8 without eye opening for at least 24 hours post-injury and age 18-65 years. Individual subject information can be found in the Supplementary Table.

MRI data for all healthy subjects and patients with TBI were acquired on a 3T Siemens Skyra scanner (Siemens Medical Solutions, Erlangen, Germany) using a 32-channel head coil. T1-weighted data were acquired using a MultiEcho Magnetization-Prepared RApid Gradient Echo (MEMP-RAGE) sequence (TR: 2530ms, TE: 1.69/3.55/5.41/7.27ms, flip angle: 7°) at 1mm isotropic spatial resolution. A 3D susceptibility-weighted imaging (SWI) sequence (TR: 30ms, TE: 20ms, flip angle: 15°) was acquired for each subject at a 0.86 x 0.86 x 1.8 mm spatial resolution.

#### 2.2.4 Alzheimer’s disease data

All T1 MRI data used in this article was obtained from the Alzheimer’s Disease Neuroimaging Initiative (ADNI) database (adni.loni.usc.edu). The ADNI was launched in 2003 as a public-private partnership, led by Principal Investigator Michael W. Weiner, MD. The original goal of ADNI was to test whether serial magnetic resonance imaging, positron emission tomography, other biological markers, and clinical and neuropsychological assessment can be combined to measure the progression of mild cognitive impairment and early AD. Specifically, we used T1 MRI images (scanned at approximately 1mm isotropic resolution) from 215 randomly-chosen subjects with AD (mean age: 75.53 ± 7.38) and 168 age-matched control subjects (mean age: 76.09 ± 5.43) and. Further information on subject and acquision information can be found at adni.loni.usc.edu. All aforementioned ADNI subjects were used in previous classification analyses (Iglesias, Augustinack, et al., 2015; Iglesias, Van Leemput, et al., 2015; Saygin et al., 2017).

#### 2.2.5 Human Connectome Project data

We used 45 subjects from the Human Connectome Project (HCP) WU-Minn 1200 subject release dataset (Van Essen et al., 2013) who underwent two separate scanning sessions (included as part of the “Retest Data” cohort) with the same scanning protocols to assess test-retest reliability. Specifically, we analyzed unprocessed T1-weighted (TR: 2400ms, TE: 2.14ms, flip angle: 8°) and T2-weighted TR: 3200ms, TE: 565ms, flip angle: 120°) MRI images scanned at 0.7mm isotropic resolution. Further information can be found at db.humanconnectome.org.

### 2.3 *Ex Vivo* manual annotation of brainstem arousal nuclei

For *ex vivo* brain specimens with corresponding histological sections (S1, S2), AAN nuclei were traced on the DWSSFP 750 µm average low-b image, which was directly used for probabilistic atlas construction, as described in section 2.4.2. Tracing on the low-b image was performed with guidance from the 7T MEF 200 µm FLASH image, which provided contrast for the boundaries of smaller AAN nuclei. AAN nuclei labels were further refined with tyrosine hydroxylase (for VTA and LC staining), tryptophan hydroxylase (for DR and MnR staining), and H&E-LFB stains (annotation of all other AAN nuclei) to accurately translate the location and morphology of the nuclei from (ground-truth) histological to low-b space. Manual tracing of AAN nuclei in *ex vivo* brain specimens without histological sections (S3-S5) was performed with guidance from the 200 µm FLASH structural images. Neuroanatomic boundaries of all AAN nuclei were cross-referenced with the Harvard Disorders of Consciousness Histopathology Collection (http://histopath.nmr.mgh.harvard.edu) and the Paxinos human brainstem atlas (Paxinos et al., 2012). Additional details regarding the anatomic locations and annotation protocol of AAN nuclei have been previously described (Edlow et al., 2024).

### 2.4 Probabilistic atlas construction and Bayesian segmentation

#### 2.4.1 Ex Vivo MRI dataset

To optimize the accuracy of segmentation, a probabilistic atlas needs to describe not only the neuroanatomical structures of interest, but also their surrounding tissue. With this purpose, we ran our sequence adaptive segmentation method (SAMSEG, (Puonti et al., 2016)) on the low-b image of the *ex vivo* scans to obtain labels for 36 different brain structures, including the whole brainstem; left and right cerebellar gray and white matter; fourth ventricle; left and right ventral diencephalon; and the left and right thalamus. After manual correction of errors made by SAMSEG, the manual segmentations of the arousal nuclei were overlaid on these automated segmentations to create composite label maps including both the brainstem nuclei of interest and surrounding structures. These composite maps were used to create the probabilistic atlas using a Bayesian technique, as described in 2.4.2 below (Figure 2). A fly-though visualization of the atlas mesh is provided in the Supplementary Video.

**Figure 1:**
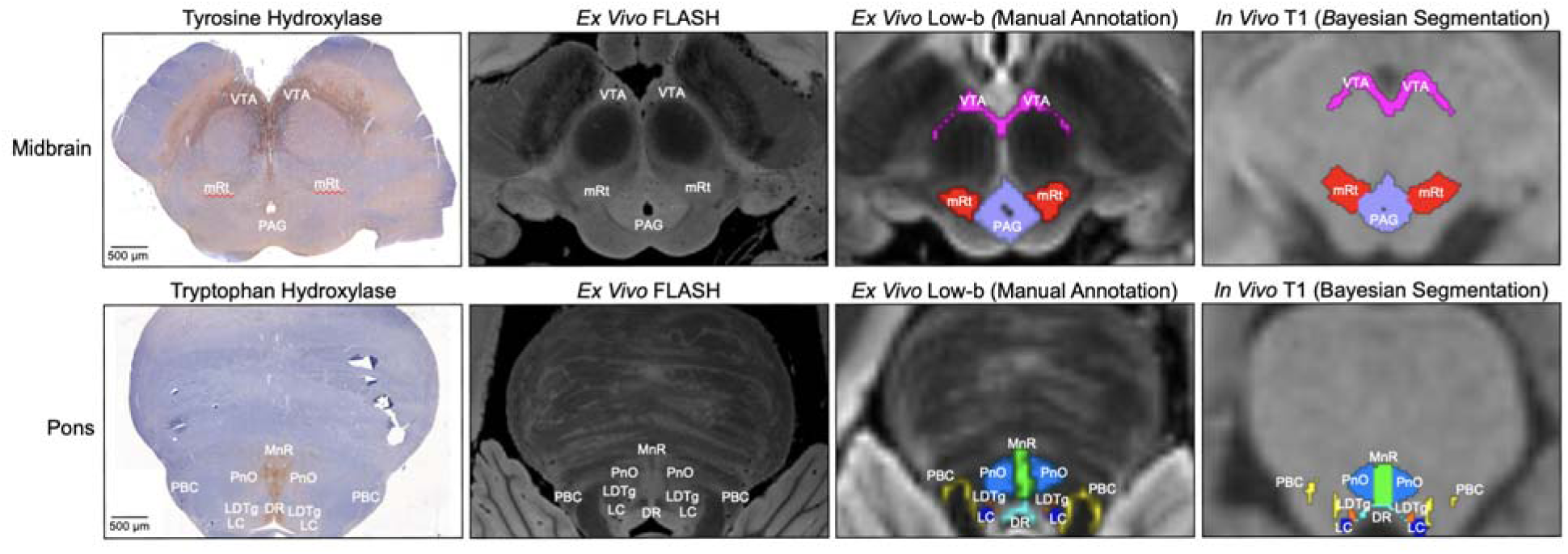
Histology-imaging correlations of AAN nuclei. Corresponding histological sections (first column) and/or 200μm FLASH images (second column) were used for manually annotating AAN nuclei in two of the five low-B diffusion MRI images from *ex vivo* brain specimens (third column). All five specimens were subsequently used for generating a probabilistic AAN atlas, which provides spatial priors for the automated Bayesian segmentation of AAN nuclei in MRI images of any contrast (fourth column).

**Figure 2:**
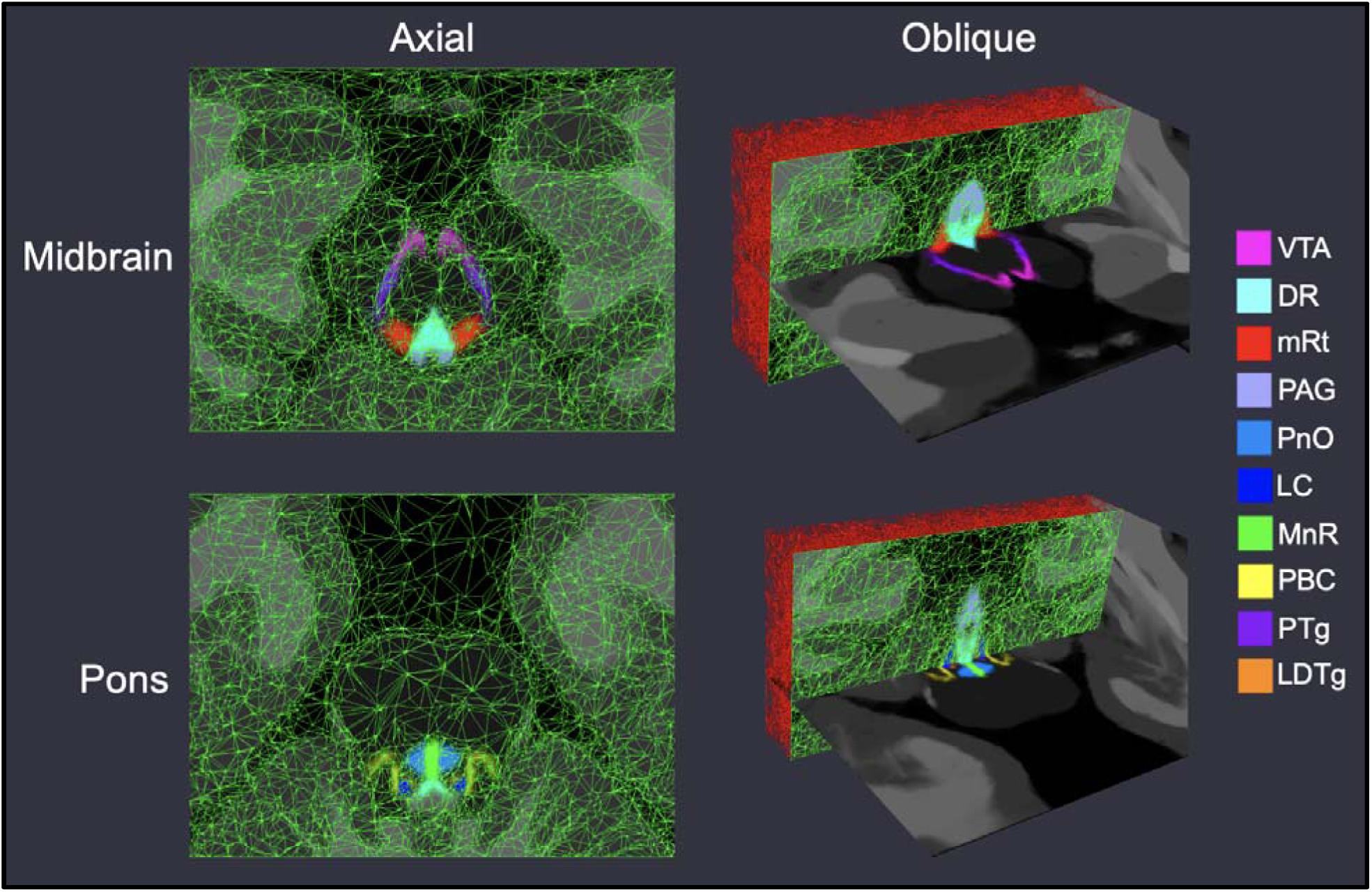
Probabilistic atlas mesh of ascending arousal network nuclei. We show axial and oblique views (with coronal mesh sections) of the adaptive probabilistic atlas mesh used to encode spatial priors for all nuclei in the midbrain (top row) and pons (bottom row). Mesh node density corresponds to the relative amount of intensity information used for atlas construction, and subsequent Bayesian segmentation. All SAMSEG-derived brain structures (including the whole-brainstem) used for atlas construction are displayed with grey intensities.

#### 2.4.2 Generative model of segmentations and atlas construction

The Bayesian segmentation framework relies on a generative model of brain MRI data, where neuroanatomy and model formation are decoupled (Ashburner & Friston, 2005; Van Leemput et al., 1999). This approach enables the use of *ex vivo* data of superior quality to model neuroanatomy through a probabilistic atlas and apply an atlas to the automated segmentation of *in vivo* scans or arbitrary MR contrast.

The generative model of Bayesian segmentation assumes that segmentations are generated by a probabilistic atlas. Here we used the representation proposed by (Van Leemput, 2009), where a probabilistic atlas in encoded as a tetrahedral mesh endowed with a deformation model. Every mesh node has an associated vector with the probabilities of the different neuroanatomical classes happening at each location, and such probabilities can be evaluated at any other location with interpolation. The forward model is as follows: if ***x***^*r*^ is reference position of the mesh nodes, a deformed position ***x*** is first obtained from the probability distribution:

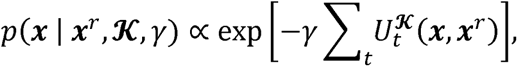

where *γ* is a scalar representing the stiffness of the deforming mesh, 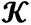 is the connectivity (topology) of the mesh, and 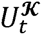 is a potential function that penalizes the deformation of the *t^th^* tetrahedron, going to infinity as the Jacobian determinant goes to zero (if the tetrahedron folds onto itself), and thus preserving the topology of the mesh (Ashburner et al., 2000). Given the deformed mesh, and the label probabilities at each node **α** = {**α**_t_}, the probability of observing class *k* at a certain voxel location j is obtained with barycentric interpolation:

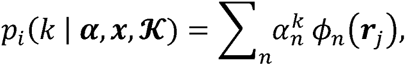

where ***r**_j_* is the spatial location of node *n* and 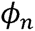 is an interpolation basis function attached to it. The generative model of segmentations is completed by assuming that a segmentation or label map *L* is obtained by sampling these label probabilities independently at each voxel location:

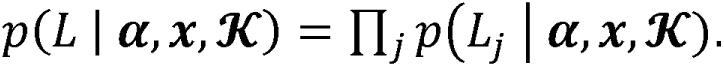

Building the atlas requires “inverting” the model with Bayesian inference, in order to estimate its parameters (reference position, label probabilities and topology) from a set of *M* example segmentations 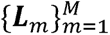. Assuming no prior knowledge on the distribution of these parameters, the problem to solve is:

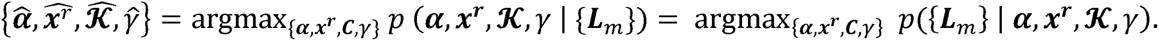

This maximization is difficult because of the need to optimize for the connectivity, which is a Bayesian model selection problem. We use a “greedy algorithm” that starts from a very dense mesh, and slowly merging tetrahedra where appropriate, using Bayesian model selection. Further details can be found in (Van Leemput, 2009).

#### 2.4.3 Segmentation as Bayesian inference

The full generative model of Bayesian segmentation combines two components: the prior, and the likelihood. The prior describes the distribution of segmentations 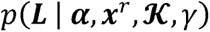, as described in Section 2.4.2 above. The likelihood describes the distribution of observed image intensities given a segmentation *L*. Here we follow the classical model of Bayesian segmentation and assume that: *(i)* each class *k* has an associated Gaussian distribution with mean *μ_k_* and variance 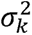; and *(ii)* the intensity of voxel *j* is an independent sample of the Gaussian distribution associated with its label *L_j_*.Therefore:

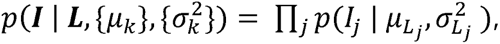

where ***I*** represents the observed image intensities, and 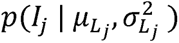 is simply the Gaussian distribution 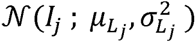.

Using Bayesian inference, segmentation within this model can be posed as the following optimization problem:

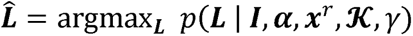

However, this requires marginalizing over model parameters, including the mesh deformation ***x***, which is intractable. Instead, the standard approximation is to compute point estimates for the parameters, and then solve the segmentation. The objective function to optimize is:

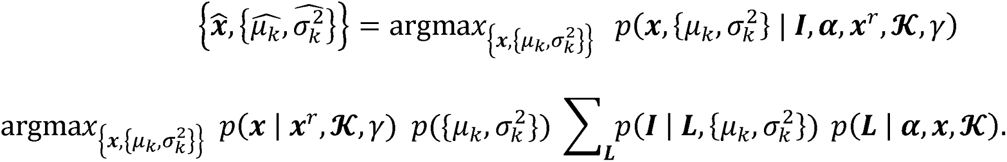

where 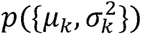 encodes prior knowledge on the Gaussian parameters, if available. Optimization is performed with a coordinate ascent strategy, alternately maximizing for the atlas deformation **x** (with the L-BFGS algorithm, (Byrd et al., 1995)) and the Gaussian parameters 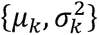 (with the Expectation Maximization algorithm (Dempster et al., 1977)). It is straightforward to show that the final segmentation, as well as the expectation of volumes of the different structures, are obtained as a byproduct of the EM algorithm. Further details can be found in (Van Leemput, 2009).

#### 2.4.4 Implementation details of segmentation method

Because linear deformation aspects were not considered in the generative model, external affine alignment is required both for atlas building and segmentation. In atlas building, we align a binary mask consisting of the whole brainstem (including arousal nuclei), left and right ventral DC and thalami to the corresponding grouping of structures in the FreeSurfer atlas, and consider only a cuboid enclosing the mask with a margin of 15 mm. In segmentation, we assume that the scan to segment has been run through the main FreeSurfer stream. Then, we can similarly co-register the same subset of structures in the FreeSurfer atlas with a binary mask including the thalami, ventral DC and brainstem, as automatically estimated by FreeSurfer.

A crucial aspect for segmentation robustness is to group structures with similar intensity profiles into superclasses. Therefore, all gray matter structures in the cerebrum (cerebral cortex, hippocampus, amygdala) share a single set of Gaussian parameters, and so do CSF structures (lateral, inferior lateral, third and fourth ventricles) and brainstem structures (brainstem, ventral DC and arousal nuclei except for the PAG, which displays some contrast and has its own Gaussian distribution). The rest of structures in the atlas have their own sets of Gaussian parameters, including the caudate nucleus, accumbens area, putamen, pallidum, thalamus, choroid plexus, cerebral white matter, cerebellar white matter, cerebellar cortex and background).

At segmentation, we also exploit the output of the main FreeSurfer stream in two ways. First, we use the coarse skull stripping provided by FreeSurfer to remove most of the extracerebral tissue. Second, we use the automated segmentation (ASEG) to inform the Gaussian parameters for each superclass (except for the PAG) as with the median intensity of the voxels within each segment using a conjugate prior. Further details can be found in (Iglesias, Van Leemput, et al., 2015).

## 3 Results

### 3.1 Segmentation accuracy for in vivo T1 scans from control and TBI subjects

We compared AAN nuclei generated with the Bayesian segmentation algorithm to manually annotated AAN segmentations in *in vivo* T1-weighted MRI scans from ten healthy control subjects and ten patients with acute severe TBI to assess segmentation performance in the setting of structural brainstem injury. We subsequently chose a subset of patients TBI who had either significant deformation of the brainstem due to increased intraventricular pressure and/or herniation, or hemorrhagic lesions in the brainstem, detected by SWI. We used SWI for annotation of the hemorrhagic lesions, as this sequence creates particularly high contrast in regions of hemorrhage that retain paramagnetic blood products such as oxy/deoxyhemoglobin, hemosiderin and methemoglobin.

Figure 3 shows the Dice coefficients and corresponding 95th-percentile Haussdorf distances (HDs) for the ten control subjects and ten deformed/lesioned brainstems from patients with severe TBI. For two binary masks *M_a_* and *M_b_*, Dice coefficients and HDs are defined as:

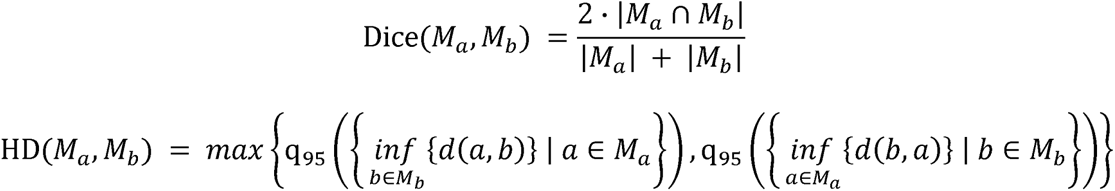

where | · | represents the cardinality (volume) of a region, *d*(· , ·) represents Euclidean distance and q_95_({·}) represents the 95th percentile (quantile) value of a set. As expected, Bayesian segmentations of nuclei with thin cross-sectional areas, mainly the LC (Dice: 0.38), LDTg (Dice: 0.19), and PBC (Dice: 0.19), displayed the lowest degrees of direct overlap with manual annotations in control subjects. The rest of the AAN nuclei displayed consistently better overlap with mean Dice coefficients over 0.5. While such degrees of overlap are low for standard segmentation algorithms, this is expected given that AAN nuclei are orders of magnitude smaller than most regions in the brain that are segmented by standard algorithms. HD, a distance metric that captures boundary precision and is less sensitive to small changes in overlap (as compared to Dice coefficients), was less than 2 mm for all AAN nuclei except for the PBC in control subjects (HD: 2.12 mm), reflecting the high precision of the algorithm. Minimal alteration in Dice coefficients and 95HD was observed for the ten TBI patients, with only slight decreases in segmentation accuracy and precision. This observation indicates that the Bayesian segmentation algorithm is, to a degree, robust to lesioning and/or deformation in the brainstem, as shown in Figure 4. Notable though, we observed poorer performance with significantly large (albeit rare) brainstem lesions, where the algorithm tends to inpatient lesioned regions as locations of proximal nuclei, such as in Figure 5.

**Figure 3.**
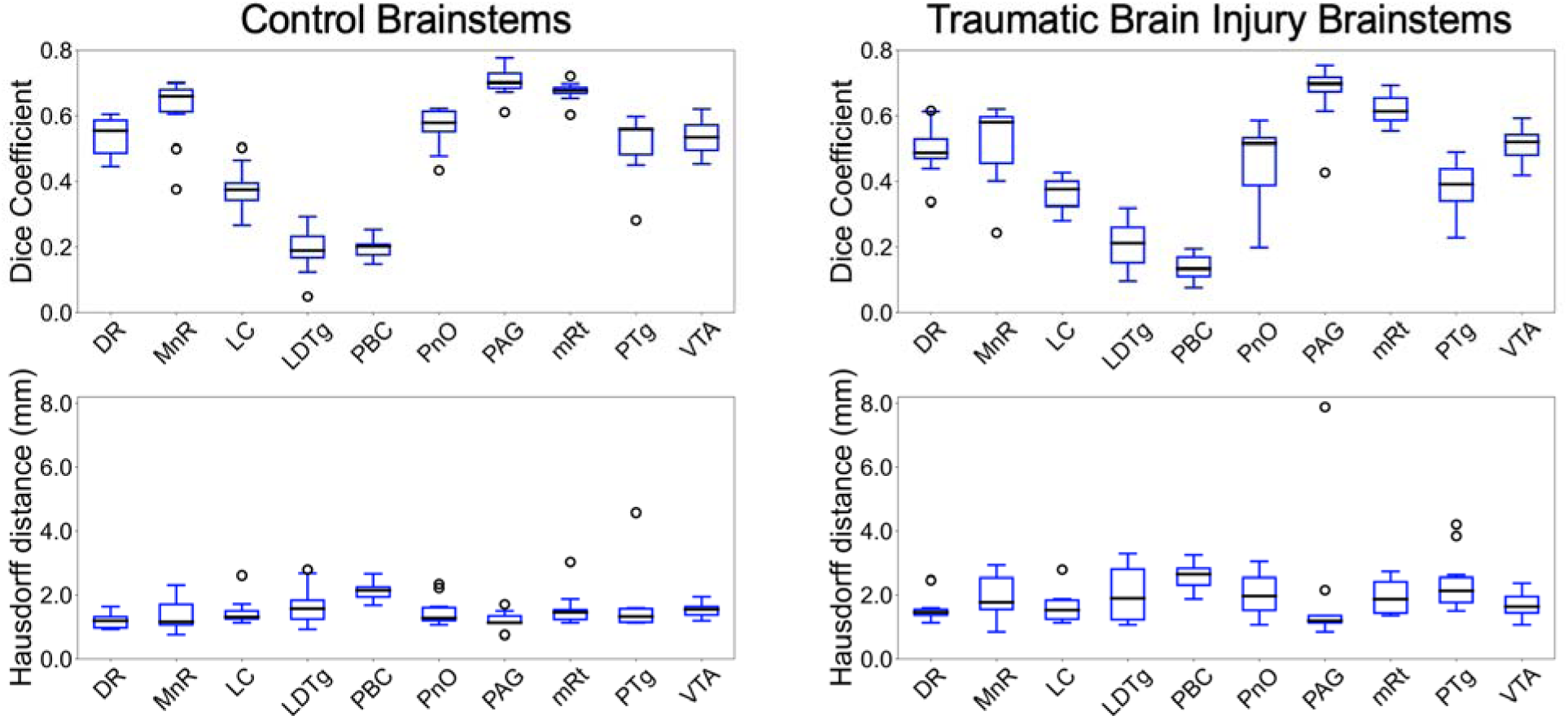
Bayesian segmentation accuracy in control and TBI T1 MRI scans. Displayed in the left panel are box plots of Dice coefficients (top) and associated 95% Hausdorff distances (bottom) from direct comparisons of automated segmentations and manual annotation of each AAN nucleus in ten T1 MRI scans from healthy control subjects. Shown in the right panel are Dice scores (top) and HD (bottom) derived from 10 Traumatic Brain Injury patients with deformed and/or lesioned brainstems. Divisions of the box plots are 25th percentile, median, and 75th percentile.

**Figure 4.**
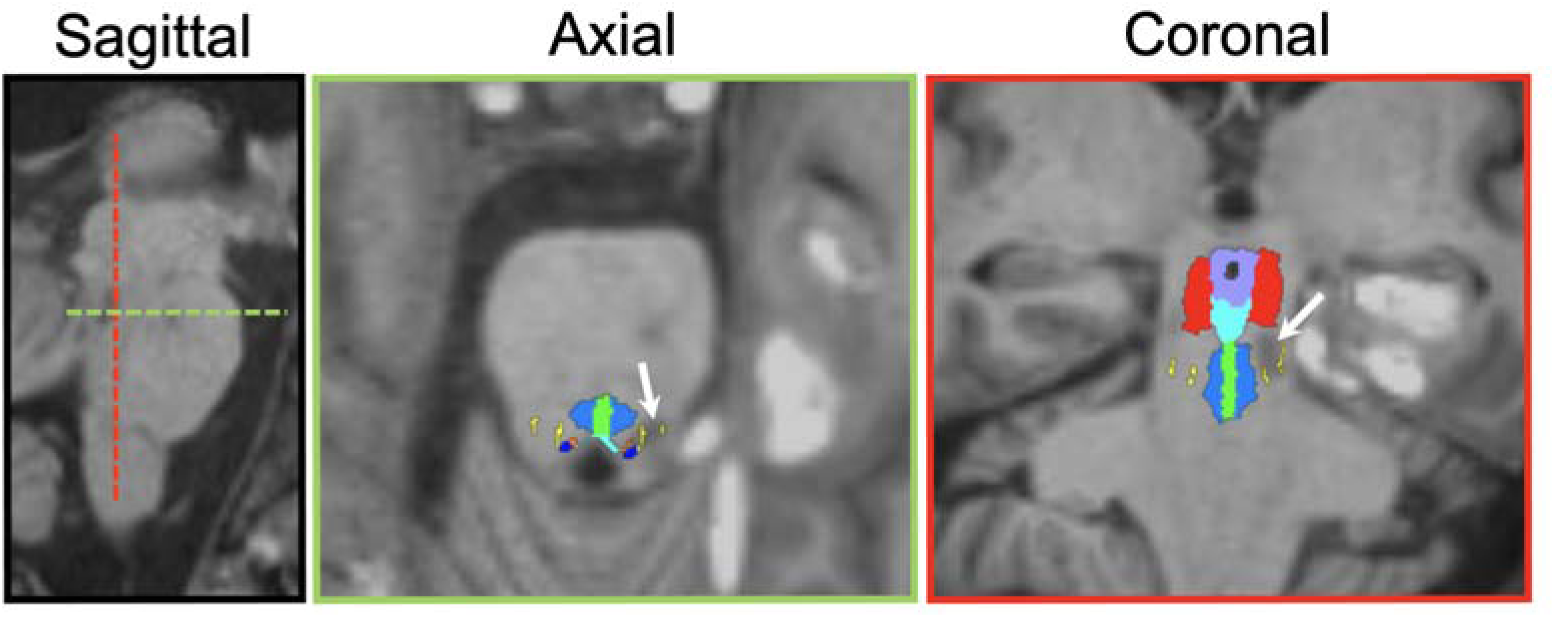
Bayesian segmentation of AAN nuclei in a lesioned and deformed brainstem. The white arrows in each panel point to the location of a hemorrhagic lesion detected by susceptibility-weighted imaging.

**Figure 5.**
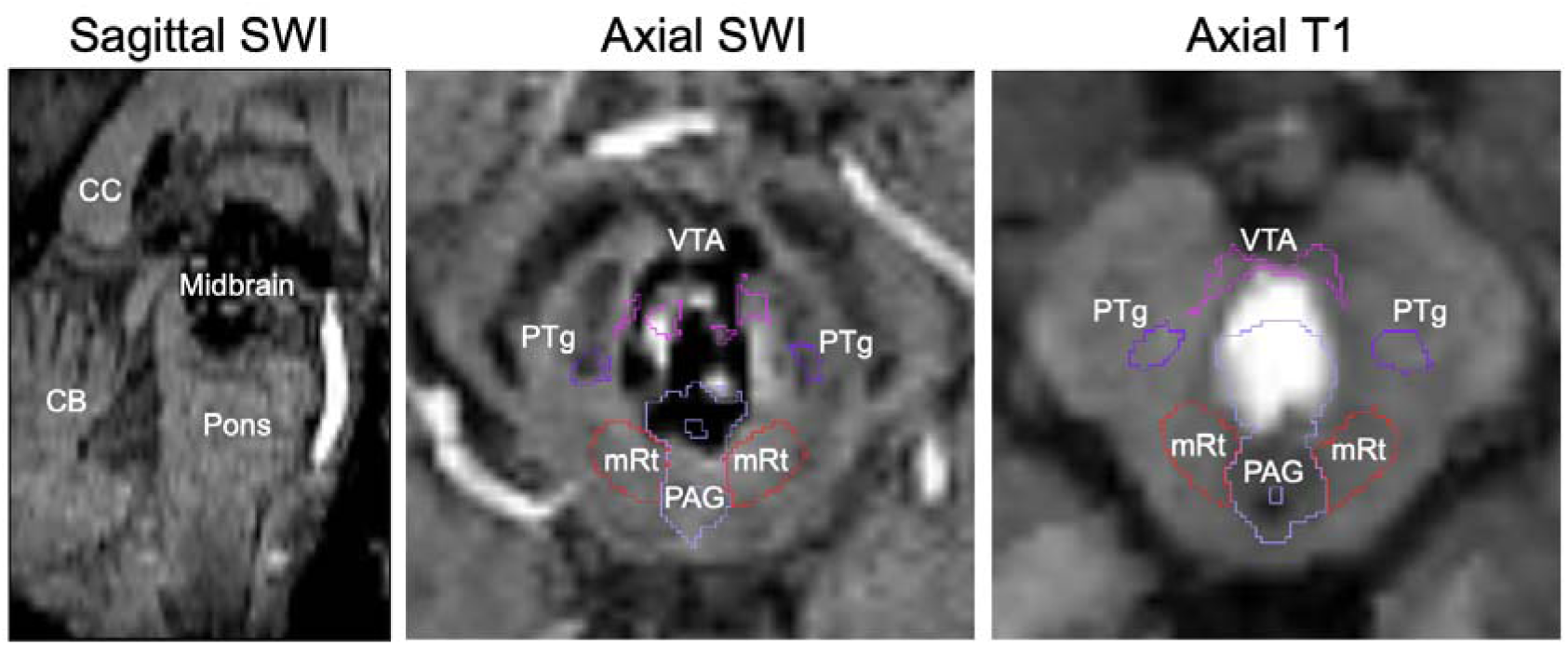
Segmentation inpainting of a large brainstem lesion. MRI scanning of patient 15 (see Supplementary Table) revealed a large hemorrhagic lesion spanning the entirety of the midbrain, as seen in the sagittal view (left), and bordering the PAG, mRt, PTg, and VTA. Automated segmentation of AAN nuclei in both the SWI and the T1 images yielded significant inpainting of the PAG, and to a lesser degree the VTA, inside of the lesion margins.

### 3.2 Test-retest analysis

We observed robust test-retest reliability for both T1 and T2 MRI scans of subjects from the HCP “retest” dataset (Figure 6). Intraclass Correlation Coefficients (ICCs) are expected to be low for segmentations of small regions that are prone to fluctuations of estimated volume, but we observed excellent reliability (ICC > 0.75) for all AAN segmentations except for the right LC and right LDTg for analysis in the same domain (i.e., T1-T1 and T2-T2). We also observed ICC>0.7 across domains (i.e., T1-T2 and T2-T1) with exception for the DR, PAG, LC and LDTg. Furthermore, while there was a small drop-off in ICC with decreasing segmentation volume, as shown in the volume-versus-ICC scatter plot (Figure 6A), this relationship was not statistically significant based on a Wald t-test for deviation from a zero-slope null hypothesis (*R* = 0.26, *p* = 0.33). This result indicates that that Bayesian AAN segmentation reliability does not decline with the spatial volume of the AAN nucleus.

**Figure 6:**
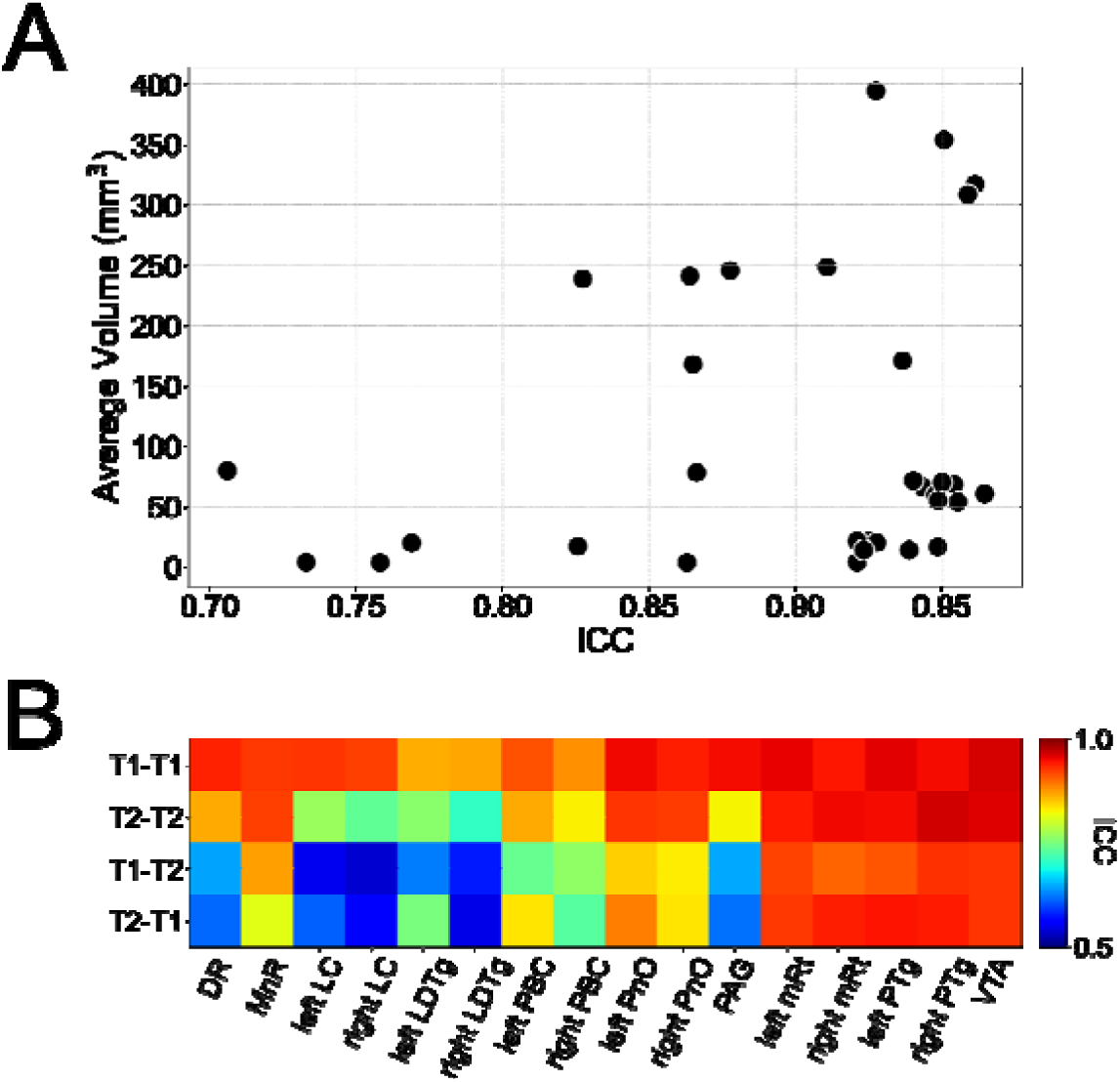
Test-retest analysis for volumes of segmented AAN nuclei. Shown in panel (A) are volumes of segmented AAN nuclei compared to their respective ICC values for T1-T1 and T2-T2 comparisons. Each scatter point corresponds to volumes are averaged amongst the entire test-retest cohorts and between the ‘test’ and the ‘retest’ scanning sessions. Only T1-T1 and T2-T2 test-retest comparisons are displayed due to redundancy of inclusion of cross-model segmentations. Shown in panel (B) are the ICC values for the segmented volume of each AAN nucleus for test-retest analysis between two MR contrasts: T1 in the test set and T1 in the retest set (T1-T1), T2 in the test set and T2 in the retest set (T2-T2), T1 in the test set and T2 in the retest set (T1-T2), and T2 in the test set and T1 in the retest set (T2-T1).

### 3.3 Classification performance in Alzheimer’s disease

To show the clinical translatability of Bayesian AAN segmentation in group studies, we assessed changes in spatial volume of each AAN nucleus in T1 MRI scans between healthy and AD subjects from the ADNI dataset. Significant decreases in the volume of the whole brainstem, particularly rostral midbrain volumes, have been reported in individuals with AD (J. H. Lee et al., 2015). To our knowledge no volumetric analysis has been performed on AAN nuclei, even though histologic alterations in several AAN nuclei, including the locus coeruleus and the raphe nuclei, have been implicated in AD progression (Chen et al., 2022; Simic et al., 2009).

For our volume classification model, we implemented a Linear Discriminant Analysis (LDA) classifier, due to its linearity and simplicity (Figure 7). This allows for the underlying volumes of AAN nuclei to more directly affect discriminatory performance, as opposed to the explicit parameterization of the classifier. We trained the LDA classifier with the volume of each segmented AAN nucleus as a separate feature with leave-one-out cross-validation. For classification analysis, we constructed a Receiver-Operating Characteristic (ROC) curve by varying the threshold of the LDA likelihood ratio. We also implemented corresponding ROC curves by varying the classification threshold for the volumes of the entire masked AAN, as well as the whole brainstem extracted from the FreeSurfer *aseg* package. For statistical comparison, we calculated Areas Under the Curve (AUCs) for each classifier, as well as paired DeLong tests (DeLong et al., 1988) between pairs of classifiers.

**Figure 7.**
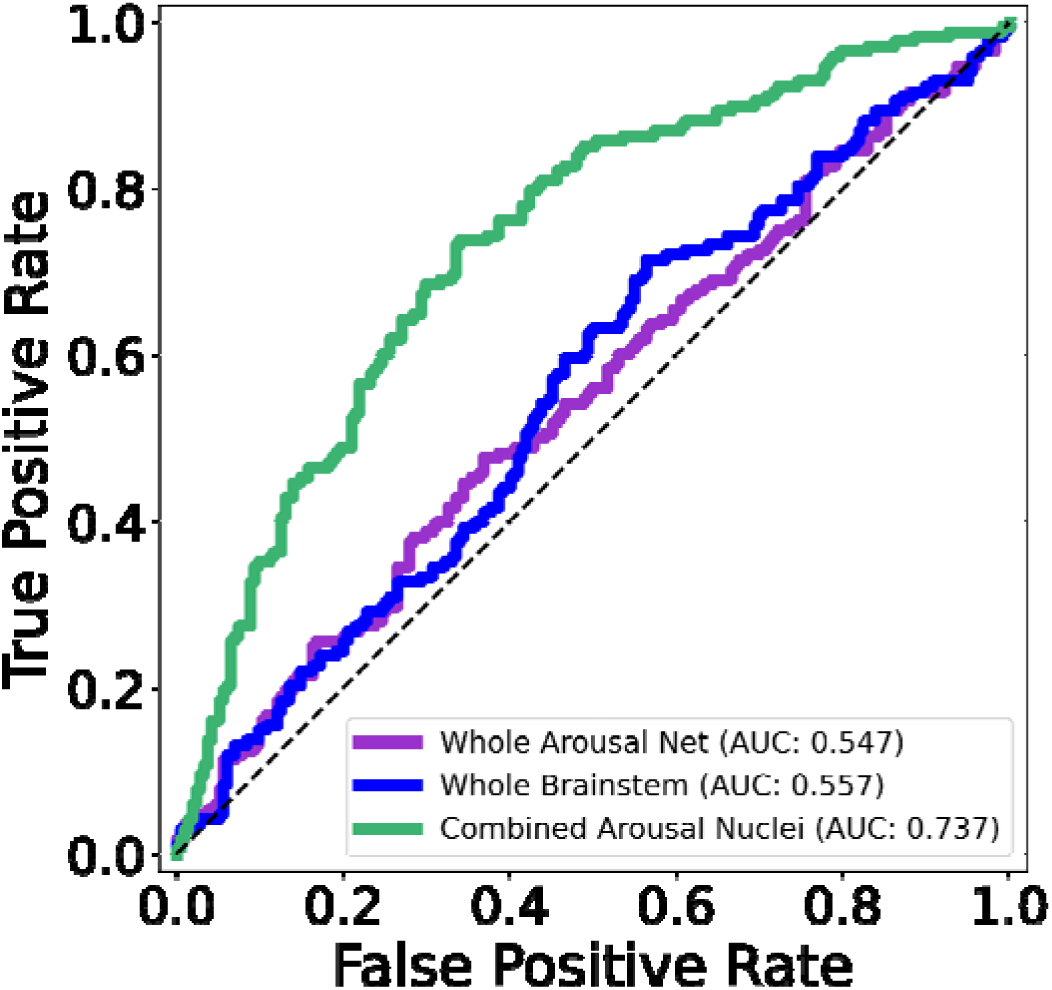
ROC analysis of AAN segmentations in Alzheimer’s Disease. Displayed in the left panel are ROC curves for a classification task (between healthy control and AD subjects in the ADNI dataset) based on a varying threshold for the volumes of a brainstem mask (blue), the segmented whole-AAN mask (purple), and the likelihood ratio of an LDA classifier trained on individual segmented AAN nuclei (green).

Classification performance based on the respective ROC curves were comparably poor for the brainstem and whole-AAN masks with AUCs of 0.58 and 0.57 respectively. Conversely, the LDA classifier built on individual AAN nuclei significantly outperformed both whole-brainstem and whole-AAN classification, with an AUC of 0.75 and a DeLong p<0.001 for comparisons with both of the aforementioned classifiers. This significant boost in classification power of the LDA classifier is most likely attributable to the individual classification power of a large subset of the AAN nuclei. Eight out of the 16 nuclei (excluding their left-right subdivisions) showed volume reduction in the AD cohort that was deemed statistically significant with Bonferroni correction (Table 3). Notably, all of the AAN nuclei with significant volume reduction have previously been shown to exhibit morphological and/or histological alterations in individuals with AD (Mufson et al., 1988; Parvizi et al., 1998, 2000, 2001).

**Table 3:**
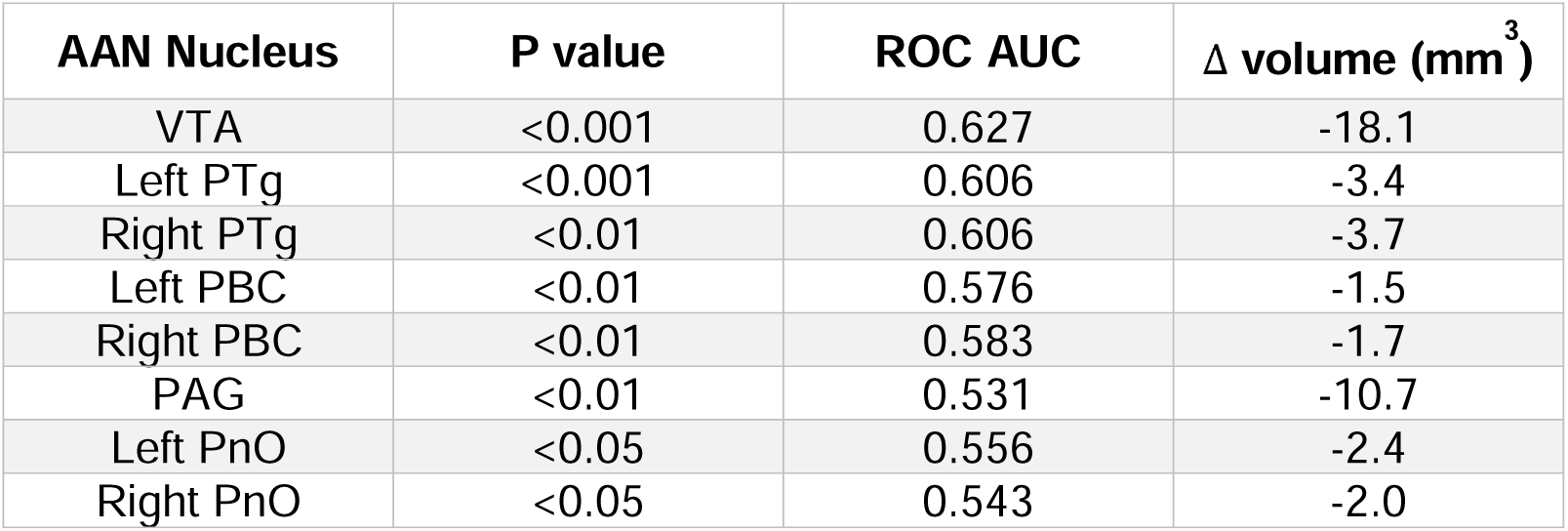
Shown are the segmented AAN nuclei which have displayed a statistically-significant reduction in volume in the AD cohort compared to healthy controls based on a Bonferroni-corrected (n=16) two-tailed Wilcoxon Rank-Sum test. The third column displays the net reduction in cohort-averaged volumes for each AAN nucleus.

| AAN Nucleus | P value | ROC AUC | $\Delta$ volume (mm<sup>3</sup>) |
| --- | --- | --- | --- |
| VTA | <0.001 | 0.627 | -18.1 |
| Left PTg | <0.001 | 0.606 | -3.4 |
| Right PTg | <0.01 | 0.606 | -3.7 |
| Left PBC | <0.01 | 0.576 | -1.5 |
| Right PBC | <0.01 | 0.583 | -1.7 |
| PAG | <0.01 | 0.531 | -10.7 |
| Left PnO | <0.05 | 0.556 | -2.4 |
| Right PnO | <0.05 | 0.543 | -2.0 |

### 3.4 Correlations with susceptibility-weighted imaging in patients with severe TBI

Fifteen of the 18 patients from the TBI dataset underwent SWI scanning (see protocol in Section 2.2.3). SWI intensities, which are sensitive to hemorrhagic lesions commonly observed in TBI (Bianciardi et al., 2021; Tao et al., 2015), were masked by the overlay of all segmented AAN nuclei (henceforth referred to as an AAN mask). The masked SWI intensities were then averaged and normalized with the average SWI intensity observed in the lateral ventricles (segmented via the FreeSurfer SynthSeg tool, and manually corrected to remove hypointense regions of hemorrhage in the cerebrospinal fluid). Averaged and normalized SWI intensities were compared to two metrics that were used to assess each patient’s level of consciousness (LoC) at the time of the MRI: the Total GCS (GCS-T) and the Total Coma Recovery Scale-Revised (CRSR-T) (Giacino et al., 2004). Assessment of both LoC metrics yielded strong positive correlations (R = 0.65 and 0.60 for GCS-T and CRSR-T correlations, respectively) (Figure 8). Both correlations were statistically significant based on a zero-slope null-hypothesis (p = 0.008 and 0.018 for GCS-T and CRSR-T correlations, respectively).

**Figure 8:**
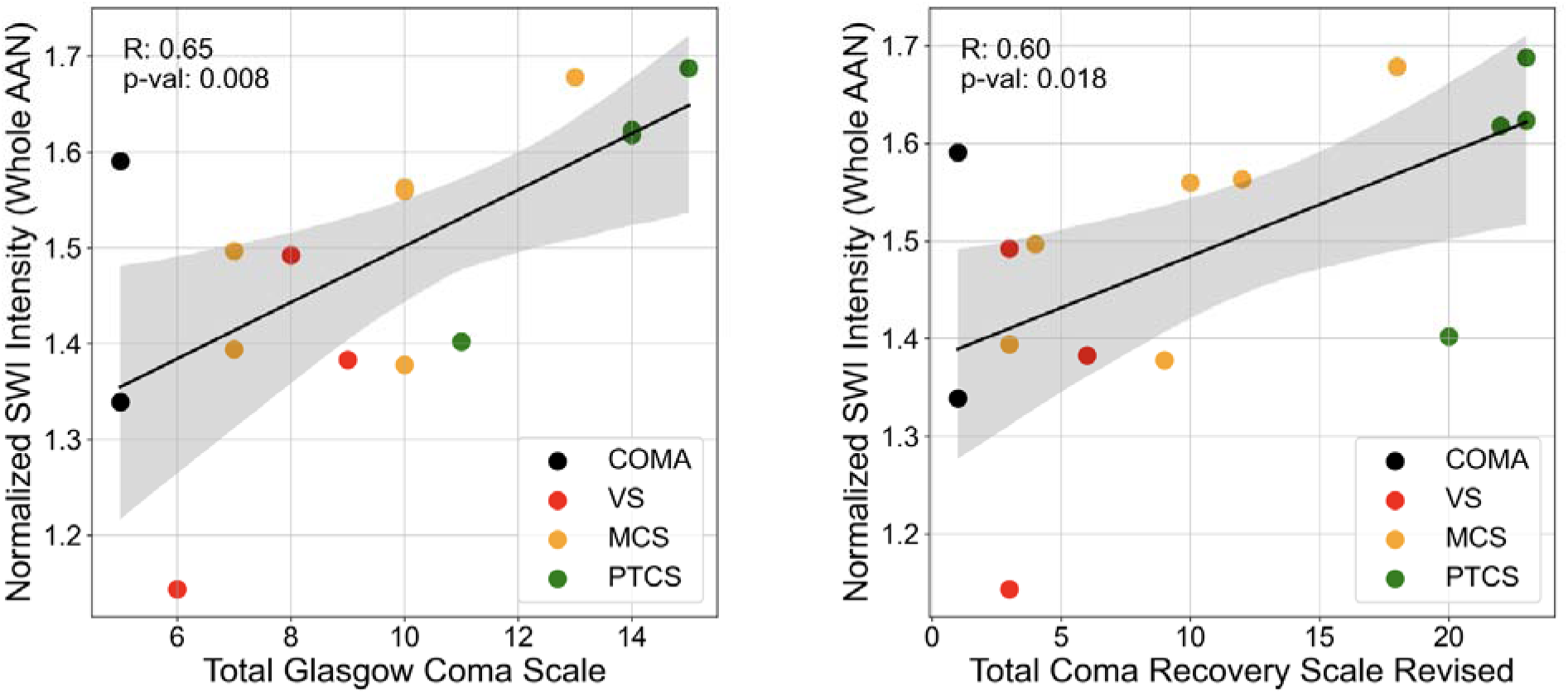
SWI AAN correlations with LoC. SWI intensities were averaged across segmented AAN masks and normalized by average SWI intensities in the lateral ventricles across all TBI patient scans. This normalized AAN SWI value was calculated for 16 acute TBI patients and correlated with their assigned LoC assessments: the GCS-T (left panel), and the CRSR-T (right panel). Coloring of the scatter points indicate each TBI patient’s general LoC classification: coma, VS: vegetative state, MCS: Minimally-Conscious State, and PTCS: Post-Traumatic Confusional State. R values are derived directly from a linear least-squares regression and p values indicate significance of deviation of the regression lines from a zero-slope based on a two-tailed Wald t-test.

## 4 Discussion

Historically, a major barrier to advancing knowledge about the brainstem’s role in the physiology of human consciousness and the pathophysiology of DoC has been the lack of tools for identifying and segmenting the tiny grey matter nuclei of the brainstem’s AAN. Here, we develop and disseminate an automated segmentation tool and a probabilistic atlas of ten AAN nuclei built from immunostaining data and meticulous manual segmentation of AAN nuclei in five ultra-high-resolution *ex vivo* MRI datasets. The AAN atlas generation process is built on a Bayesian framework, which can segment AAN nuclei in any MRI modality. We validated this process with comparisons to *in vivo* manual annotations, as well as assessment of test-retest reliability. We demonstrate the clinical translatability of the method by assessing volumetric changes of AAN nuclei in AD, and by correlating AAN SWI intensities with behavioral measures of consciousness in severe TBI.

The Bayesian AAN segmentation tool developed here builds upon recent efforts by progressing from segmentation of medullary, pontine, and midbrain subregions (Iglesias, Van Leemput, et al., 2015) to segmentation of individual AAN nuclei. We demonstrate the accuracy of the AAN segmentation tool in manually traced *in vivo* T1 data with guidance from a brainstem atlas. This manually traced dataset consisted of a 10-subject control dataset as well as a 10-patient TBI dataset where brainstems were either lesioned or deformed due to mass effect from hemorrhagic and/or edematous intracranial lesions. Dice coefficients for larger AAN nuclei were all greater than 0.5. While modest compared to previously published Dice coefficients for segmentations of larger brain structures (Fischl et al., 2002; Iglesias, Van Leemput, et al., 2015; Wasserthal et al., 2018), the reported AAN Dice coefficients are expected given the relatively small size of AAN nuclei. This challenge is especially relevant to the smaller nuclei (e.g., LC, LDTg, and PBC), whose cross-sections can be as thin as one voxel (approximately 0.4 mm). Our reported AAN Dice coefficients are comparable to algorithms which segment other small brain regions such as hypothalamic and thalamic nuclei (Billot et al., 2020; Tregidgo et al., 2023). Notably, there was less variability in HDs (a metric that is largely invariant to ROI size) between segmentations of AAN nuclei and their corresponding manual annotations, with all average distances being below 2 mm. The low HDs imply that our automated segmentations retain spatial specificity for all AAN nuclei regardless of size.

The robustness of the Bayesian AAN segmentation tool is further demonstrated by the test-retest reliability analyses. We compared the spatial volumes of segmented AAN nuclei in T1 and T2 sequences of HCP subjects who underwent to separate scanning sessions. Test-retest ICC scores for AAN volumes derived from the same sequence (T1-T1, and T2-T2) were all greater than 0.8, with the exception of LC and LDTg. Test-retest ICC scores for AAN nuclei volumes derived from alternating sequences (T1-T2, and T2-T1) were greater than 0.7, with the exception of LC, LDTg, DR, PAG. For the majority of AAN nuclei, the high repeatability of volume measurement reflects that segmentations are leveraging actual MR contrast as opposed to noise. Notably, each of the nuclei with low ICCs (LC, LDTg, DR and PAG) are positioned proximal to tissue-CSF boundaries, which are prone to high noise levels. Brainstem-CSF boundaries, especially the dorsal-brainstem-fourth-ventricle border, have significantly higher signal-to-noise ratios due to both partial-volume averaging and CSF pulsatility effects, which have been observed in dorsal brainstem nuclei such as the PAG and DR in prior literature (Sclocco et al., 2018). These results suggest that further optimization of the segmentation tool is needed in future studies, and that analyses of AAN nuclei located at brainstem-CSF boundaries should be interpreted with caution.

To demonstrate the potential translational utility of the automated AAN segmentation tool, we performed proof-of-principle analyses of AAN segmentation in patients with AD and severe TBI. In AD subjects, the segmentation tool detected differences in AAN volumes in patients with AD, as compared to healthy control subjects. Neuropathologic changes in AAN nuclei have been reported in pathology studies of patients with AD (Rüb et al., 2016; Uematsu et al., 2018) but MRI studies of changes in the AAN are scarce (Galgani et al., 2023; Miyoshi et al., 2013; Takahashi et al., 2015), likely due to the difficulty of both manual and automated AAN segmentation. Whereas classification of volumetric differences was similar for the brainstem and whole-AAN masks, there was a significant boost in classification performance for the LDA classifier trained on individual AAN nuclei, with an AUC increase of 0.18 (LDA classifier versus whole-AAN mask). In each AAN nucleus with a statistically significant change in volume, we observed a reduction in volume in the AD patient group. This volume loss is consistent with prior MRI studies, which reported volumetric reduction in the midbrain and pontine tegmentum where most AAN nuclei are located (Ji et al., 2020; J. H. Lee et al., 2015). The VTA, which contains dopaminergic neurons that widely project to and modulate numerous cortical regions (Morales & Margolis, 2017), displayed the most significant volumetric decrease in AD patients. This observation is supported by previous imaging and neuropathology studies which found that AD progression correlates with volume loss and dopamine neuron degeneration in the VTA (Bozzali et al., 2016, 2019; De Marco & Venneri, 2018; Gibb et al., 1989).

Finally, we provide evidence for the robustness and potential clinical utility of Bayesian AAN segmentation using a non-standard MRI sequence for the assessment of severe TBI. Specifically, we show an association between the number of traumatic microhemorrhages detected by SWI within the AAN and the LoC of each patient. Brainstem microhemorrhages, which are caused by traumatic shearing of arterioles and venules, are a hallmark finding of traumatic DoC because of their association with axonal shearing and disconnection of the neural networks that support consciousness (Edlow et al., 2013). Microhemorrhages within AAN nuclei appear to have particular prognostic relevance (Bianciardi et al., 2021; Izzy et al., 2017). We observed a correlation between the normalized average SWI intensity of the AAN mask and LoC in a cohort of acute TBI patients, such that patients with lower LoC displayed lower SWI intensities (i.e., more microhemorrhages). This association between SWI intensity and LoC was observed with two behavioral assessments scores used to evaluate LoC in clinical practice: the GCS (R=0.65) and the CRS-R (R=0.60).

Our findings in patients with severe TBI complement those of a previously published SWI analysis of this same cohort, with the key distinction that the prior analysis required laborious and time-consuming manual segmentation, whereas the present analysis utilized a rapid, robust automated segmentation tool. The prior study showed associations between the total SWI lesion volume in the AAN and the duration of unresponsiveness in TBI patients (Bianciardi et al., 2021). Our analysis of SWI hypo-intensities in Bayesian-segmented AAN nuclei extends this study in two ways: by showing strong associations between SWI contrast in the AAN and LoC in the acute phase of severe TBI, and by demonstrating the feasibility of an automated approach to to measurement of AAN micro haemorrhages in TBI patients based on imaging biomarkers.

Several limitations should be considered when interpreting the Bayesian segmentation methods, as well as the experimental results. First, the segmentation tool has the potential to produce irregular AAN segmentations in highly deformed and/or heavily lesioned brainstem regions, such as inpainting large (hyperintense) hemorrhagic lesions in T1 MRI scans. This irregularity likely reflects a limitation of the probabilistic atlas design, where there is a trade-off between accommodating high tissue distortion and extreme tissue intensities during atlas building but losing regularization strength for anatomically plausible atlas meshing. Furthermore, the likelihood for each tissue class in the probabilistic atlas is represented by a single Gaussian distribution, which limits the scope of model fitting in lesioned tissue. Future directions will involve incorporating distributions that can better capture intensity variations in the presence of lesions, and to evaluate the utility of convolutional neural networks (CNN) as an alternative AAN segmentation method for lesioned or deformed brainstems. CNNs may provide enhanced flexibility and ability to capture disease-specific contrast patterns, which are infeasible to encode in a probabilistic atlas. Furthermore, the implementation of a CNN could improve segmentation of small AAN nuclei and modelling of partial-volume and CSF pulsatility effects, thereby enhancing the segmentation accuracy of AAN nuclei in close proximity to the fourth ventricle and cerebral aqueduct.

Second, we note the small sample size for both the *ex vivo* data used to fit the probabilistic atlas, and the *in vivo* data used for comparisons of Bayesian segmentations to manual labels (with only a single rater performing *in vivo* annotations). Although only five *ex vivo* specimens were used for atlas building, low sample numbers for the formation of generative models (i.e., the probabilistic AAN atlas) are common and often beneficial, as to prevent model underfitting (Ng & Jordan, 2001). Furthermore, the relative rarity of normative *ex vivo* brains without pathology, coupled with the time commitment necessary for brain processing, makes the formation of a high-sample number atlas model challenging.

Third, all manual annotations of AAN nuclei in both *ex vivo* and *in vivo* MRI data was performed by a single rater. While all label annotations were confirmed by a neuropathologist (H.C.K.) and neurologist (B.L.E.) with expertise in brainstem anatomy, single-rater annotation may have introduced bias in the assessment of segmentation performance. This limitation can be addressed in future studies with a second rater re-annotating the *in vivo* dataset and performing inter-rater variability analysis.

Finally, our analysis is currently limited to T1, T2, and SWI MRI scans with isotropic spatial resolutions at or smaller than 1mm. While these are commonly used sequences in clinical imaging, further assessment of the generalizability of this segmentation method should be performed in other MRI domains. These domains can include lower-resolution diffusion and/or functional MRI sequences, emerging ultra-low-field sequences that are used in the intensive care unit (Sheth et al., 2021), and novel sequences that enhance contrast of deep-brain structures which are of interest to brainstem imaging (Sclocco et al., 2018).

## 5 Conclusion

We present a probabilistic atlas of brainstem arousal nuclei within the AAN, a subcortical network whose connections are believed to be critical for recovery of consciousness in patients with coma (Brown et al., 2010; Edlow et al., 2021; Schiff & Plum, 2000). We generated the AAN atlas from *ex vivo* MRI with histological guidance, which allowed for highly accurate manual delineation of nuclei with boundaries that lacked MRI contrast. This probabilistic atlas serves as a backbone for a Bayesian segmentation method that allows for automated delineation of AAN nuclei in MRI scans of any contrast. We show that the Bayesian method produces accurate AAN segmentations in both healthy and lesioned/structurally-deformed brainstems and is highly reliable across multiple MRI contrasts in a test-retest analysis. Furthermore, morphology and intensity information from AAN nuclei have the potential to serve as imaging-based biomarkers for AD and severe TBI. Future directions to improve the proposed AAN segmentation method will include utilizing multiple contrasts, which would be especially beneficial for nuclei with faint contrast boundaries, as well as the use of a CNN to supplant or replace Bayesian inference. Future analysis will include the investigation of imaging-based AAN biomarkers in longitudinal analysis of coma recovery, as well the study of brainstem-centric neurodegenerative disorders. We release the automated tool to advance the study of human brainstem anatomy in consciousness and its disorders.

## Data Availability

Representative ex vivo data that are used for method generation and accuracy analysis are openly available in OpenNeuro at (https://openneuro.org/datasets/ds004640/versions/1.0.4), reference number (ds004640). Corresponding histology and immunostaining data are available at (histopath.nmr.mgh.harvard.edu). The segmentation algorithm and probabilistic version of the Harvard Ascending Arousal Network Atlas are being released via github and are integrated into the FreeSurfer software platform (https://surfer.nmr.mgh.harvard.edu/fswiki/AANSegment).

https://histopath.nmr.mgh.harvard.edu/images/?page=images&selectionType=collection&selectionId=45

## Acknowledgments

This study was supported by the NIH National Institute of Neurological Disorders and Stroke (R21NS109627, RF1NS115268, U01NS086625, U54NS115322, R01NS128961, U24NS135561, U01NS132181, U01NS137484), NIH Director’s Office (DP2 HD101400), NIH National Institute of Biomedical Imaging and Bioengineering (T32EB001680), NIH National Institute of Mental Health (RF1MH123195), NIH National Institute on Aging (R01AG070988), United States Department of Defense (W81XWH2210999), European Research Council Starting Grant (677697), Alzheimer’s Research UK Interdisciplinary grant (ARUK-IRG2019A-003), MIT-Takeda Fellowship, Rappaport Foundation, James S. McDonnell Foundation, MIT/MGH Brain Arousal State Control Innovation Center (BASCIC) project, and the Chen Institute MGH Research Scholar Award. We thank Kathryn Regan and Veranex Inc. for assistance with immunostaining and histology.

Data were provided in part by the Human Connectome Project, WU-Minn Consortium (Principal Investigators: David Van Essen and Kamil Ugurbil; 1U54MH091657) funded by the 16 NIH Institutes and Centers that support the NIH Blueprint for Neuroscience Research; and by the McDonnell Center for Systems Neuroscience at Washington University.

Collection and sharing of ADNI data were performed by the Alzheimer’s Disease Neuroimaging Initiative (ADNI), which is funded by the National Institute on Aging (National Institutes of Health Grant U19AG024904). The grantee organization is the Northern California Institute for Research and Education. In the past, ADNI has also received funding from the National Institute of Biomedical Imaging and Bioengineering, the Canadian Institutes of Health Research, and private sector contributions through the Foundation for the National Institutes of Health (FNIH) including generous contributions from the following: AbbVie, Alzheimer’s Association; Alzheimer’s Drug Discovery Foundation; Araclon Biotech; BioClinica, Inc.; Biogen; Bristol-Myers Squibb Company; CereSpir, Inc.; Cogstate; Eisai Inc.; Elan Pharmaceuticals, Inc.; Eli Lilly and Company; EuroImmun; F. Hoffmann-La Roche Ltd and its affiliated company Genentech, Inc.; Fujirebio; GE Healthcare; IXICO Ltd.; Janssen Alzheimer Immunotherapy Research & Development, LLC.; Johnson & Johnson Pharmaceutical Research & Development LLC.; Lumosity; Lundbeck; Merck & Co., Inc.; Meso Scale Diagnostics, LLC.; NeuroRx Research; Neurotrack Technologies; Novartis Pharmaceuticals Corporation; Pfizer Inc.; Piramal Imaging; Servier; Takeda Pharmaceutical Company; and Transition Therapeutics.

## References

Adil, S. M., Calabrese, E., Charalambous, L. T., Cook, J. J., Rahimpour, S., Atik, A. F., Cofer, G. P., Parente, B. A., Johnson, G. A., Lad, S. P., & White, L. E. (2021). A high-resolution interactive atlas of the human brainstem using magnetic resonance imaging. NeuroImage, 237, 118135. 10.1016/j.neuroimage.2021.118135

Ashburner, J., Andersson, J. L., & Friston, K. J. (2000). Image registration using a symmetric prior--in three dimensions. Human Brain Mapping, 9(4), 212–225. 10.1002/(sici)1097-0193(200004)9:4<212::aid-hbm3>3.0.co;2-#

Ashburner, J., & Friston, K. J. (2005). Unified segmentation. NeuroImage, 26(3), 839–851. 10.1016/j.neuroimage.2005.02.018

Augustinack, J. C., van der Kouwe, A. J. W., Blackwell, M. L., Salat, D. H., Wiggins, C. J., Frosch, M. P., Wiggins, G. C., Potthast, A., Wald, L. L., & Fischl, B. R. (2005). Detection of entorhinal layer II using Tesla magnetic resonance imaging. Annals of Neurology, 57(4), 489–494. 10.1002/ana.20426

Beissner, F., Schumann, A., Brunn, F., Eisenträger, D., & Bär, K.-J. (2014). Advances in functional magnetic resonance imaging of the human brainstem. NeuroImage, 86, 91–98. 10.1016/j.neuroimage.2013.07.081

Bianciardi, M. (2021). Brainstem Navigator [Tool/Resource.] (https://www.nitrc.org/doi/landing_page.php?table=groups&id=1551). Washington: NITRC.

Bianciardi, M., Izzy, S., Rosen, B. R., Wald, L. L., & Edlow, B. L. (2021). Location of Subcortical Microbleeds and Recovery of Consciousness After Severe Traumatic Brain Injury. Neurology, 97(2). 10.1212/WNL.0000000000012192

Bianciardi, M., Toschi, N., Eichner, C., Polimeni, J. R., Setsompop, K., Brown, E. N., Hämäläinen, M. S., Rosen, B. R., & Wald, L. L. (2016). In vivo functional connectome of human brainstem nuclei of the ascending arousal, autonomic, and motor systems by high spatial resolution 7-Tesla fMRI. *Magma (New York*, N.Y.), 29(3), 451–462. 10.1007/s10334-016-0546-3

Billot, B., Bocchetta, M., Todd, E., Dalca, A. V., Rohrer, J. D., & Iglesias, J. E. (2020). Automated segmentation of the hypothalamus and associated subunits in brain MRI. NeuroImage, 223, 117287. 10.1016/j.neuroimage.2020.117287

Bodien, Y. G., Chatelle, C., & Edlow, B. L. (2017). Functional Networks in Disorders of Consciousness. Seminars in Neurology, 37(5), 485–502. 10.1055/s-0037-1607310

Boes, A. D., Prasad, S., Liu, H., Liu, Q., Pascual-Leone, A., Caviness Jr, V. S., & Fox, M. D. (2015). Network localization of neurological symptoms from focal brain lesions. Brain : A Journal of Neurology, 138(Pt 10), 3061–3075. 10.1093/brain/awv228

Bondiau, P.-Y., Malandain, G., Chanalet, S., Marcy, P.-Y., Habrand, J.-L., Fauchon, F., Paquis, P., Courdi, A., Commowick, O., Rutten, I., & Ayache, N. (2005). Atlas-based automatic segmentation of MR images: Validation study on the brainstem in radiotherapy context. International Journal of Radiation Oncology*Biology*Physics, 61(1), 289–298. 10.1016/j.ijrobp.2004.08.055

Bozzali, M., D’Amelio, M., & Serra, L. (2019). Ventral tegmental area disruption in Alzheimer’s disease. Aging, 11(5), 1325–1326. 10.18632/aging.101852

Bozzali, M., Serra, L., & Cercignani, M. (2016). Quantitative MRI to understand Alzheimer’s disease pathophysiology. Current Opinion in Neurology, 29(4), 437–444. 10.1097/WCO.0000000000000345

Brown, E. N., Lydic, R., & Schiff, N. D. (2010). General anesthesia, sleep, and coma. The New England Journal of Medicine, 363(27), 2638–2650. 10.1056/NEJMra0808281

Buckner, R. L., & DiNicola, L. M. (2019). The brain’s default network: updated anatomy, physiology and evolving insights. Nature Reviews Neuroscience, 20(10), 593–608. 10.1038/s41583-019-0212-7

Byrd, R. H., Lu, P., Nocedal, J., & Zhu, C. (1995). A Limited Memory Algorithm for Bound Constrained Optimization. SIAM Journal on Scientific Computing, 16(5), 1190–1208. 10.1137/0916069

Chen, Y., Chen, T., & Hou, R. (2022). Locus coeruleus in the pathogenesis of Alzheimer’s disease: A systematic review. Alzheimer’s & Dementia: Translational Research & Clinical Interventions, 8(1). 10.1002/trc2.12257

Cole, M. W., Bassett, D. S., Power, J. D., Braver, T. S., & Petersen, S. E. (2014). Intrinsic and task-evoked network architectures of the human brain. Neuron, 83(1), 238–251. 10.1016/j.neuron.2014.05.014

De Marco, M., & Venneri, A. (2018). Volume and Connectivity of the Ventral Tegmental Area are Linked to Neurocognitive Signatures of Alzheimer’s Disease in Humans. Journal of Alzheimer’s Disease, 63(1), 167–180. 10.3233/JAD-171018

DeLong, E. R., DeLong, D. M., & Clarke-Pearson, D. L. (1988). Comparing the Areas under Two or More Correlated Receiver Operating Characteristic Curves: A Nonparametric Approach. Biometrics, 44(3), 837. 10.2307/2531595

Demertzi, A., Tagliazucchi, E., Dehaene, S., Deco, G., Barttfeld, P., Raimondo, F., Martial, C., Fernández-Espejo, D., Rohaut, B., Voss, H. U., Schiff, N. D., Owen, A. M., Laureys, S., Naccache, L., & Sitt, J. D. (2019). Human consciousness is supported by dynamic complex patterns of brain signal coordination. Science Advances, 5(2). 10.1126/sciadv.aat7603

Dempster, A. P., Laird, N. M., & Rubin, D. B. (1977). Maximum Likelihood from Incomplete Data Via the *EM* Algorithm. Journal of the Royal Statistical Society Series B: Statistical Methodology, 39(1), 1–22. 10.1111/j.2517-6161.1977.tb01600.x

Edlow, B. L., Chatelle, C., Spencer, C. A., Chu, C. J., Bodien, Y. G., O’Connor, K. L., Hirschberg, R. E., Hochberg, L. R., Giacino, J. T., Rosenthal, E. S., & Wu, O. (2017). Early detection of consciousness in patients with acute severe traumatic brain injury. Brain, 140(9), 2399–2414. 10.1093/brain/awx176

Edlow, B. L., Claassen, J., Schiff, N. D., & Greer, D. M. (2021). Recovery from disorders of consciousness: mechanisms, prognosis and emerging therapies. Nature Reviews Neurology, 17(3), 135–156. 10.1038/s41582-020-00428-x

Edlow, B. L., Haynes, R. L., Takahashi, E., Klein, J. P., Cummings, P., Benner, T., Greer, D. M., Greenberg, S. M., Wu, O., Kinney, H. C., & Folkerth, R. D. (2013). Disconnection of the ascending arousal system in traumatic coma. Journal of Neuropathology and Experimental Neurology, 72(6), 505–523. 10.1097/NEN.0b013e3182945bf6

Edlow, B. L., Mareyam, A., Horn, A., Polimeni, J. R., Witzel, T., Tisdall, M. D., Augustinack, J. C., Stockmann, J. P., Diamond, B. R., Stevens, A., Tirrell, L. S., Folkerth, R. D., Wald, L. L., Fischl, B., & van der Kouwe, A. (2019). 7 Tesla MRI of the ex vivo human brain at 100 micron resolution. Scientific Data, 6(1), 244. 10.1038/s41597-019-0254-8

Edlow, B. L., McNab, J. A., Witzel, T., & Kinney, H. C. (2016). The Structural Connectome of the Human Central Homeostatic Network. Brain Connectivity, 6(3), 187–200. 10.1089/brain.2015.0378

Edlow, B. L., Olchanyi, M., Freeman, H. J., Li, J., Maffei, C., Snider, S. B., Zöllei, L., Iglesias, J. E., Augustinack, J., Bodien, Y. G., Haynes, R. L., Greve, D. N., Diamond, B. R., Stevens, A., Giacino, J. T., Destrieux, C., van der Kouwe, A., Brown, E. N., Folkerth, R. D., … Kinney, H. C. (2024). Multimodal MRI reveals brainstem connections that sustain wakefulness in human consciousness. Science Translational Medicine, 16(745). 10.1126/scitranslmed.adj4303

Edlow, B. L., Takahashi, E., Wu, O., Benner, T., Dai, G., Bu, L., Grant, P. E., Greer, D. M., Greenberg, S. M., Kinney, H. C., & Folkerth, R. D. (2012). Neuroanatomic Connectivity of the Human Ascending Arousal System Critical to Consciousness and Its Disorders. Journal of Neuropathology & Experimental Neurology, 71(6), 531–546. 10.1097/NEN.0b013e3182588293

Fischer, D. B., Boes, A. D., Demertzi, A., Evrard, H. C., Laureys, S., Edlow, B. L., Liu, H., Saper, C. B., Pascual-Leone, A., Fox, M. D., & Geerling, J. C. (2016). A human brain network derived from coma-causing brainstem lesions. Neurology, 87(23), 2427–2434. 10.1212/WNL.0000000000003404

Fischl, B. (2012). FreeSurfer. NeuroImage, 62(2), 774–781. 10.1016/j.neuroimage.2012.01.021

Fischl, B., Salat, D. H., Busa, E., Albert, M., Dieterich, M., Haselgrove, C., van der Kouwe, A., Killiany, R., Kennedy, D., Klaveness, S., Montillo, A., Makris, N., Rosen, B., & Dale, A. M. (2002). Whole Brain Segmentation. Neuron, 33(3), 341–355. 10.1016/s0896-6273(02)00569-x

Fischl, B., Salat, D. H., van der Kouwe, A. J. W., Makris, N., Ségonne, F., Quinn, B. T., & Dale, A. M. (2004). Sequence-independent segmentation of magnetic resonance images. NeuroImage, 23, S69–S84. 10.1016/j.neuroimage.2004.07.016

Fox, M. D., Snyder, A. Z., Vincent, J. L., Corbetta, M., Van Essen, D. C., & Raichle, M. E. (2005). The human brain is intrinsically organized into dynamic, anticorrelated functional networks. Proceedings of the National Academy of Sciences of the United States of America, 102(27), 9673–9678. 10.1073/pnas.0504136102

Galgani, A., Lombardo, F., Martini, N., Vergallo, A., Bastiani, L., Hampel, H., Hlavata, H., Baldacci, F., Tognoni, G., De Marchi, D., Ghicopulos, I., De Cori, S., Biagioni, F., Busceti, C. L., Ceravolo, R., Bonuccelli, U., Chiappino, D., Siciliano, G., Fornai, F., … Giorgi, F. S. (2023). Magnetic resonance imaging Locus Coeruleus abnormality in amnestic Mild Cognitive Impairment is associated with future progression to dementia. European Journal of Neurology, 30(1), 32–46. 10.1111/ene.15556

Giacino, J. T., Kalmar, K., & Whyte, J. (2004). The JFK Coma Recovery Scale-Revised: Measurement characteristics and diagnostic utility11No commercial party having a direct financial interest in the results of the research supporting this article has or will confer a benefit upon the authors or upon any organization with which the authors are associated. Archives of Physical Medicine and Rehabilitation, 85(12), 2020–2029. 10.1016/j.apmr.2004.02.033

Gibb, W. R., Mountjoy, C. Q., Mann, D. M., & Lees, A. J. (1989). The substantia nigra and ventral tegmental area in Alzheimer’s disease and Down’s syndrome. *Journal of Neurology*, Neurosurgery & Psychiatry, 52(2), 193–200. 10.1136/jnnp.52.2.193

Glasser, M. F., Coalson, T. S., Robinson, E. C., Hacker, C. D., Harwell, J., Yacoub, E., Ugurbil, K., Andersson, J., Beckmann, C. F., Jenkinson, M., Smith, S. M., & Van Essen, D. C. (2016). A multi-modal parcellation of human cerebral cortex. Nature, 536(7615), 171–178. 10.1038/nature18933

Heckemann, R. A., Hajnal, J. V, Aljabar, P., Rueckert, D., & Hammers, A. (2006). Automatic anatomical brain MRI segmentation combining label propagation and decision fusion. NeuroImage, 33(1), 115–126. 10.1016/j.neuroimage.2006.05.061

Horn, A., Ostwald, D., Reisert, M., & Blankenburg, F. (2014). The structural–functional connectome and the default mode network of the human brain. NeuroImage, 102, 142–151. 10.1016/j.neuroimage.2013.09.069

Huang, C., Huang, L., Wang, Y., Li, X., Ren, L., Gu, X., Kang, L., Guo, L., Liu, M., Zhou, X., Luo, J., Huang, Z., Tu, S., Zhao, Y., Chen, L., Xu, D., Li, Y., Li, C., Peng, L., … Cao, B. (2021). 6-month consequences of COVID-19 in patients discharged from hospital: a cohort study. *Lancet (London*, England), 397(10270), 220–232. 10.1016/S0140-6736(20)32656-8

Iglesias, J. E., Augustinack, J. C., Nguyen, K., Player, C. M., Player, A., Wright, M., Roy, N., Frosch, M. P., McKee, A. C., Wald, L. L., Fischl, B., Van Leemput, K., & Initiative, A. D. N. (2015). A computational atlas of the hippocampal formation using ex vivo, ultra-high resolution MRI: Application to adaptive segmentation of in vivo MRI. NeuroImage, 115, 117–137. 10.1016/j.neuroimage.2015.04.042

Iglesias, J. E., Insausti, R., Lerma-Usabiaga, G., Bocchetta, M., Van Leemput, K., Greve, D. N., van der Kouwe, A., Initiative, A. D. N., Fischl, B., Caballero-Gaudes, C., & Paz-Alonso, P. M. (2018). A probabilistic atlas of the human thalamic nuclei combining ex vivo MRI and histology. NeuroImage, 183, 314–326. 10.1016/j.neuroimage.2018.08.012

Iglesias, J. E., Van Leemput, K., Bhatt, P., Casillas, C., Dutt, S., Schuff, N., Truran-Sacrey, D., Boxer, A., Fischl, B., & Initiative, A. D. N. (2015). Bayesian segmentation of brainstem structures in MRI. NeuroImage, 113, 184–195. 10.1016/j.neuroimage.2015.02.065

Izzy, S., Mazwi, N. L., Martinez, S., Spencer, C. A., Klein, J. P., Parikh, G., Glenn, M. B., Greenberg, S. M., Greer, D. M., Wu, O., & Edlow, B. L. (2017). Revisiting Grade 3 Diffuse Axonal Injury: Not All Brainstem Microbleeds are Prognostically Equal. Neurocritical Care, 27(2), 199–207. 10.1007/s12028-017-0399-2

Ji, X., Wang, H., Zhu, M., He, Y., Zhang, H., Chen, X., Gao, W., & Fu, Y. (2020). Brainstem atrophy in the early stage of Alzheimer’s disease: a voxel-based morphometry study. Brain Imaging and Behavior, 15(1), 49–59. 10.1007/s11682-019-00231-3

Jiann-Der Lee, Neng-Wei Wang, Chung-Hsien Huang, Li-Chang Liu, & Chin-Song Lu. (2005). A Segmentation Scheme of Brainstem and Cerebellum using Scale-Based Fuzzy Connectedness and Deformable Contour Model. 2005 IEEE Engineering in Medicine and Biology 27th Annual Conference, 459–462. 10.1109/IEMBS.2005.1616446

Kinney, H. C., & Haynes, R. L. (2019). The Serotonin Brainstem Hypothesis for the Sudden Infant Death Syndrome. Journal of Neuropathology and Experimental Neurology, 78(9), 765–779. 10.1093/jnen/nlz062

Lambert, C., Lutti, A., Helms, G., Frackowiak, R., & Ashburner, J. (2013). Multiparametric brainstem segmentation using a modified multivariate mixture of Gaussians. NeuroImage. Clinical, 2, 684–694. 10.1016/j.nicl.2013.04.017

Lechanoine, F., Jacquesson, T., Beaujoin, J., Serres, B., Mohammadi, M., Planty-Bonjour, A., Andersson, F., Poupon, F., Poupon, C., & Destrieux, C. (2021). WIKIBrainStem: An online atlas to manually segment the human brainstem at the mesoscopic scale from ultrahigh field MRI. NeuroImage, 236, 118080. 10.1016/j.neuroimage.2021.118080

Lee, J. H., Ryan, J., Andreescu, C., Aizenstein, H., & Lim, H. K. (2015). Brainstem morphological changes in Alzheimer’s disease. NeuroReport, 26(7), 411–415. 10.1097/WNR.0000000000000362

Lee, J., Tseng, Y., Liu, L., Huang, C., Lee, S., Wu, C., & Chen, J. (2007). A 2-D Automatic Segmentation Scheme for Brainstem and Cerebellum Regions in Brain MR Imaging. Fourth International Conference on Fuzzy Systems and Knowledge Discovery (FSKD 2007), 270–274. 10.1109/FSKD.2007.2

Li, J., Curley, W. H., Guerin, B., Dougherty, D. D., Dalca, A. V., Fischl, B., Horn, A., & Edlow, B. L. (2021). Mapping the subcortical connectivity of the human default mode network. NeuroImage, 245, 118758. 10.1016/j.neuroimage.2021.118758

McNab, J. A., Jbabdi, S., Deoni, S. C. L., Douaud, G., Behrens, T. E. J., & Miller, K. L. (2009). High resolution diffusion-weighted imaging in fixed human brain using diffusion-weighted steady state free precession. NeuroImage, 46(3), 775–785. 10.1016/j.neuroimage.2009.01.008

Medaglia, J. D., Lynall, M.-E., & Bassett, D. S. (2015). Cognitive Network Neuroscience. Journal of Cognitive Neuroscience, 27(8), 1471–1491. 10.1162/jocn_a_00810

Miyoshi, F., Ogawa, T., Kitao, S. -i., Kitayama, M., Shinohara, Y., Takasugi, M., Fujii, S., & Kaminou, T. (2013). Evaluation of Parkinson Disease and Alzheimer Disease with the Use of Neuromelanin MR Imaging and 123 I-Metaiodobenzylguanidine Scintigraphy. American Journal of Neuroradiology, 34(11), 2113–2118. 10.3174/ajnr.A3567

Morales, M., & Margolis, E. B. (2017). Ventral tegmental area: cellular heterogeneity, connectivity and behaviour. Nature Reviews Neuroscience, 18(2), 73–85. 10.1038/nrn.2016.165

Mufson, E. J., Mash, D. C., & Hersh, L. B. (1988). Neurofibrillary tangles in cholinergic pedunculopontine neurons in Alzheimer’s disease. Annals of Neurology, 24(5), 623–629. 10.1002/ana.410240506

Ng, A., & Jordan, M. (2001). On Discriminative vs. Generative Classifiers: A comparison of logistic regression and naive Bayes. In T. Dietterich, S. Becker, & Z. Ghahramani (Eds.), Advances in Neural Information Processing Systems (Vol. 14). MIT Press. https://proceedings.neurips.cc/paper_files/paper/2001/file/7b7a53e239400a13bd6be6c91c4f6c4e-Paper.pdf

Nigro, S., Cerasa, A., Zito, G., Perrotta, P., Chiaravalloti, F., Donzuso, G., Fera, F., Bilotta, E., Pantano, P., Quattrone, A., & Initiative, A. D. N. (2014). Fully automated segmentation of the pons and midbrain using human T1 MR brain images. PloS One, 9(1), e85618–e85618. 10.1371/journal.pone.0085618

Parvizi, J. (2001). Consciousness and the brainstem. Cognition, 79(1–2), 135–160. 10.1016/s0010-0277(00)00127-x

Parvizi, J., Van Hoesen, G. W., & Damasio, A. (1998). Severe pathological changes of parabrachial nucleus in Alzheimer’s disease. NeuroReport, 9(18), 4151–4154. 10.1097/00001756-199812210-00028

Parvizi, J., Van Hoesen, G. W., & Damasio, A. (2000). Selective pathological changes of the periaqueductal gray matter in Alzheimer’s disease. Annals of Neurology, 48(3), 344–353.

Parvizi, J., Van Hoesen, G. W., & Damasio, A. (2001). The selective vulnerability of brainstem nuclei to Alzheimer’s disease. Annals of Neurology, 49(1), 53–66. 10.1002/1531-8249(200101)49:1<53::AID-ANA30>3.0.CO;2-Q

Patenaude, B., Smith, S. M., Kennedy, D. N., & Jenkinson, M. (2011). A Bayesian model of shape and appearance for subcortical brain segmentation. NeuroImage, 56(3), 907–922. 10.1016/j.neuroimage.2011.02.046

Paxinos, G., Xu-Feng, H., Sengul, G., & Watson, C. (2012). Organization of Brainstem Nuclei. In The Human Nervous System (pp. 260–327). Elsevier. 10.1016/B978-0-12-374236-0.10008-2

Pohl, K. M., Fisher, J., Grimson, W. E. L., Kikinis, R., & Wells, W. M. (2006). A Bayesian model for joint segmentation and registration. NeuroImage, 31(1), 228–239. 10.1016/j.neuroimage.2005.11.044

Puonti, O., Iglesias, J. E., & Van Leemput, K. (2016). Fast and sequence-adaptive whole-brain segmentation using parametric Bayesian modeling. NeuroImage, 143, 235–249. 10.1016/j.neuroimage.2016.09.011

Rüb, U., Stratmann, K., Heinsen, H., Del Turco, D., Seidel, K., den Dunnen, W., & Korf, H.-W. (2016). The Brainstem Tau Cytoskeletal Pathology of Alzheimer’s Disease: A Brief Historical Overview and Description of its Anatomical Distribution Pattern, Evolutional Features, Pathogenetic and Clinical Relevance. Current Alzheimer Research, 13(10), 1178–1197. 10.2174/1567205013666160606100509

Saygin, Z. M., Kliemann, D., Iglesias, J. E., van der Kouwe, A. J. W., Boyd, E., Reuter, M., Stevens, A., Van Leemput, K., McKee, A., Frosch, M. P., Fischl, B., Augustinack, J. C., & Initiative, A. D. N. (2017). High-resolution magnetic resonance imaging reveals nuclei of the human amygdala: manual segmentation to automatic atlas. NeuroImage, 155, 370–382. 10.1016/j.neuroimage.2017.04.046

Schiff, N. D., & Plum, F. (2000). The Role of Arousal and “Gating” Systems in the Neurology of Impaired Consciousness. Journal of Clinical Neurophysiology, 17(5), 438–452. 10.1097/00004691-200009000-00002

Sclocco, R., Beissner, F., Bianciardi, M., Polimeni, J. R., & Napadow, V. (2018). Challenges and opportunities for brainstem neuroimaging with ultrahigh field MRI. NeuroImage, 168, 412–426. 10.1016/j.neuroimage.2017.02.052

Sheth, K. N., Mazurek, M. H., Yuen, M. M., Cahn, B. A., Shah, J. T., Ward, A., Kim, J. A., Gilmore, E. J., Falcone, G. J., Petersen, N., Gobeske, K. T., Kaddouh, F., Hwang, D. Y., Schindler, J., Sansing, L., Matouk, C., Rothberg, J., Sze, G., Siner, J., … Kimberly, W. T. (2021). Assessment of Brain Injury Using Portable, Low-Field Magnetic Resonance Imaging at the Bedside of Critically Ill Patients. JAMA Neurology, 78(1), 41. 10.1001/jamaneurol.2020.3263

Simic, G., Stanic, G., Mladinov, M., Jovanov-Milosevic, N., Kostovic, I., & Hof, P. R. (2009). Does Alzheimer’s disease begin in the brainstem? Neuropathology and Applied Neurobiology, 35(6), 532–554. 10.1111/j.1365-2990.2009.01038.x

Smith, S. M., Jenkinson, M., Woolrich, M. W., Beckmann, C. F., Behrens, T. E. J., Johansen-Berg, H., Bannister, P. R., De Luca, M., Drobnjak, I., Flitney, D. E., Niazy, R. K., Saunders, J., Vickers, J., Zhang, Y., De Stefano, N., Brady, J. M., & Matthews, P. M. (2004). Advances in functional and structural MR image analysis and implementation as FSL. NeuroImage, 23, S208–S219. 10.1016/j.neuroimage.2004.07.051

Snider, S. B., Hsu, J., Darby, R. R., Cooke, D., Fischer, D., Cohen, A. L., Grafman, J. H., & Fox, M. D. (2020). Cortical lesions causing loss of consciousness are anticorrelated with the dorsal brainstem. Human Brain Mapping, 41(6), 1520–1531. 10.1002/hbm.24892

Sporns, O., Tononi, G., & Kötter, R. (2005). The human connectome: A structural description of the human brain. PLoS Computational Biology, 1(4), e42–e42. 10.1371/journal.pcbi.0010042

Takahashi, J., Shibata, T., Sasaki, M., Kudo, M., Yanezawa, H., Obara, S., Kudo, K., Ito, K., Yamashita, F., & Terayama, Y. (2015). Detection of changes in the locus coeruleus in patients with mild cognitive impairment and <SCP>A</SCP> lzheimer’s disease: High-resolution fast spin-echo <SCP>T</SCP> 1-weighted imaging. Geriatrics & Gerontology International, 15(3), 334–340. 10.1111/ggi.12280

Tao, J., Zhang, W., Wang, D., Jiang, C., Wang, H., Li, W., Ji, W., & Wang, Q. (2015). Susceptibility weighted imaging in the evaluation of hemorrhagic diffuse axonal injury. Neural Regeneration Research, 10(11), 1879. 10.4103/1673-5374.170322

Teasdale, G., & Jennett, B. (1974). ASSESSMENT OF COMA AND IMPAIRED CONSCIOUSNESS. The Lancet, 304(7872), 81–84. 10.1016/S0140-6736(74)91639-0

Tregidgo, H. F. J., Soskic, S., Althonayan, J., Maffei, C., Van Leemput, K., Golland, P., Insausti, R., Lerma-Usabiaga, G., Caballero-Gaudes, C., Paz-Alonso, P. M., Yendiki, A., Alexander, D. C., Bocchetta, M., Rohrer, J. D., Iglesias, J. E., & Initiative, A. D. N. (2023). Accurate Bayesian segmentation of thalamic nuclei using diffusion MRI and an improved histological atlas. NeuroImage, 274, 120129. 10.1016/j.neuroimage.2023.120129

Uematsu, M., Nakamura, A., Ebashi, M., Hirokawa, K., Takahashi, R., & Uchihara, T. (2018). Brainstem tau pathology in Alzheimer’s disease is characterized by increase of three repeat tau and independent of amyloid β. Acta Neuropathologica Communications, 6(1), 1. 10.1186/s40478-017-0501-1

Valenza, G., Sclocco, R., Duggento, A., Passamonti, L., Napadow, V., Barbieri, R., & Toschi, N. (2019). The central autonomic network at rest: Uncovering functional MRI correlates of time-varying autonomic outflow. NeuroImage, 197, 383–390. 10.1016/j.neuroimage.2019.04.075

Van Essen, D. C., Smith, S. M., Barch, D. M., Behrens, T. E. J., Yacoub, E., & Ugurbil, K. (2013). The WU-Minn Human Connectome Project: An overview. NeuroImage, 80, 62–79. 10.1016/j.neuroimage.2013.05.041

Van Leemput, K. (2009). Encoding Probabilistic Brain Atlases Using Bayesian Inference. IEEE Transactions on Medical Imaging, 28(6), 822–837. 10.1109/TMI.2008.2010434

Van Leemput, K., Maes, F., Vandermeulen, D., & Suetens, P. (1999). Automated model-based tissue classification of MR images of the brain. IEEE Transactions on Medical Imaging, 18(10), 897–908. 10.1109/42.811270

Wasserthal, J., Neher, P., & Maier-Hein, K. H. (2018). TractSeg - Fast and accurate white matter tract segmentation. NeuroImage, 183, 239–253. 10.1016/j.neuroimage.2018.07.070

Wells, W. M., Grimson, W. E. L., Kikinis, R., & Jolesz, F. A. (1996). Adaptive segmentation of MRI data. IEEE Transactions on Medical Imaging, 15(4), 429–442. 10.1109/42.511747

Yendiki, A., Aggarwal, M., Axer, M., Howard, A. F. D., van Walsum, A.-M. van C., & Haber, S. N. (2022). Post mortem mapping of connectional anatomy for the validation of diffusion MRI. NeuroImage, 256, 119146. 10.1016/j.neuroimage.2022.119146

Yeo, B. T. T., Krienen, F. M., Sepulcre, J., Sabuncu, M. R., Lashkari, D., Hollinshead, M., Roffman, J. L., Smoller, J. W., Zöllei, L., Polimeni, J. R., Fischl, B., Liu, H., & Buckner, R. L. (2011). The organization of the human cerebral cortex estimated by intrinsic functional connectivity. Journal of Neurophysiology, 106(3), 1125–1165. 10.1152/jn.00338.2011

