## Supporting Material for "Histology-guided MRI segmentation of brainstem nuclei critical to consciousness"

** co-senior authors*

| Patient ID | Age  Range | Sex | Cause of TBI | Duration of coma (d) | Day of MRI scanning | Level of consciousness at MRI | GSC-T at MRI | CRSR-T at MRI |
| --- | --- | --- | --- | --- | --- | --- | --- | --- |
| 1 | 26-30 | M | MVA | 1 | 16 | PTCS | 15 | 23 |
| 2 | 21-25 | M | Ped vs car | 1 | 1 | MCS- | 7 | 4 |
| 3 | 16-20 | F | MVA | 5 | 3 | Coma | 5 | 1 |
| 4 | 16-20 | M | Fall | 1 | 17 | PTCS | 14 | 23 |
| 5 | 31-35 | M | Fall | 6 | 15 | VS | 7 | 3 |
| 6 | 26-30 | F | MVA | 2 | 7 | VS | 9 | 6 |
| 7 | 41-45 | M | MVA | 1 | 13 | MCS+ | 13 | 18 |
| 8 | 31-35 | M | Fall | 1 | 8 | PTCS | 11 | 20 |
| 9 | 31-35 | M | Ped vs car | 1 | 11 | MCS+ | 10 | 9 |
| 10 | 21-25 | M | Assault | 1 | 12 | MCS- | 10 | 10 |
| 11 | 21-25 | F | Ped vs car | 1 | 14 | PTCS | 14 | 22 |
| 12 | 26-30 | F | Fall | 13 | 8 | Coma | 5 | 1 |
| 13 | 16-20 | M | Fall | 1 | 4 | MCS+ | 10 | 12 |
| 14 | 51-55 | M | Ped vs car | 8 | 8 | VS | 6 | 3 |
| 15 | 26-30 | M | Ped vs car | 3 | 7 | MCS- | 7 | 3 |
| 16 | 31-35 | M | Fall | 3 | 3 | MCS+ | 10 | 12 |
| 17 | 26-30 | F | Ped vs car | 4 | 12 | VS | 8 | 3 |
| 18 | 26-30 | M | Fall | 1 | 28 | PTCS | 11 | 18 |

**Supplementary Table.** Demographic and clinical data for patients with traumatic brain injury (TBI). MVA: Motor Vehicle Accident; Ped vs car: Pedestrian versus car; PTCS: Post-Traumatic Confusional State; MCS+: Minimally Conscious State with evidence of language function; MCS-: Minimally Conscious State without evidence of language function; VS: vegetative state; GCS-T: Total Glasgow Coma Scale score; CRSR-T: Total Coma Recovery Scale-Revised score. Further study and subject information can be found in [Edlow et. al., *Brain* 2017].

**Supplementary Video.** Fly-through visualization of the adaptive probabilistic atlas mesh that encodes locations and intensity distributions of ascending arousal network nuclei in the brainstem. The mesh axis is displayed in the coronal plane. VTA: Ventral Tegmental Area; LC: Locus Coeruleus; DR: Dorsal Raphe; MnR: Median Raphe; mRt: Midbrain Reticular Formation; PBC: Parabrachial Complex; PAG: Periaqueductal Gray; PTg: Pedunculotegmental Nucleus; PnO: Pontis Oralis; LDTg: Laterodorsal Tegmental Nucleus.
